# An autosomal dominant cardiac arrhythmia syndrome, ST Depression Syndrome, is caused by the *de novo* creation of a cardiomyocyte enhancer

**DOI:** 10.1101/2024.08.20.24312115

**Authors:** Carin P. de Villiers, Damien J. Downes, Anuj Goel, Alistair T. Pagnamenta, Elizabeth Ormondroyd, Alexander J. Sparrow, Svanhild Nornes, Edoardo Giacopuzzi, Phalguni Rath, Ben Davies, Ron Schwessinger, Matthew E. Gosden, Robert A. Beagrie, Duncan Parkes, Rob Hastings, Stefano Lise, Silvia Salatino, Hannah Roberts, Maria Lopopolo, Carika Weldon, Amy Trebes, The WGS500 consortium, David Buck, Jenny C. Taylor, Charles Redwood, Edward Rowland, Dushen Tharmaratnam, Graham Stuart, Pier D. Lambiase, Sarah De Val, Jim R. Hughes, Hugh Watkins

**Affiliations:** Division of Cardiovascular Medicine, Radcliffe Department of Medicine, University of Oxford, Oxford, UK; MRC Molecular Haematology Unit, MRC Weatherall Institute of Molecular Medicine, Radcliffe Department of Medicine, University of Oxford, Oxford, UK; Centre for Human Genetics, University of Oxford, Oxford, UK; Ludwig Institute for Cancer Research Ltd, Nuffield Department of Medicine, University of Oxford, Oxford, UK; Department of Physiology, Anatomy and Genetics, University of Oxford, Oxford, UK; Human Technopole, Milan, Italy; MRC WIMM Centre for Computational Biology, MRC Weatherall Institute of Molecular Medicine, Radcliffe Department of Medicine, University of Oxford, Oxford, UK; Laboratory of Gene Regulation, MRC Weatherall Institute of Molecular Medicine, Radcliffe Department of Medicine, University of Oxford, Oxford, UK; University College London Cancer Institute, London, UK; St Bartholomew’s Hospital, Barts Health NHS Trust, London, UK and UCL Institute of Cardiovascular Science, University College London, London, UK; Royal Devon University Healthcare NHS Foundation Trust; Bristol Heart Institute, Bristol, United Kingdom

**Keywords:** Enhancer, KCNB1, ECG, ST-segment, Arrhythmia, Mendelian disease, gene regulation, machine learning

## Abstract

A substantial proportion of mutations underlying rare Mendelian diseases remain unknown, potentially because they lie in the non-coding genome. Here, we report the mapping of the causal mutation of an autosomal dominant cardiac arrhythmia syndrome, ST Depression Syndrome, which is associated with widespread ST-depression on the electrocardiogram together with risk of sudden death and heart failure, to the non-coding region of the *KCNB1* locus. Using genetic linkage analysis, we narrowed the associated region to 1cM of the genome and then with a genome editing approach, we show that the mutation, a small complex insertion-deletion, generates a *de novo* gain-of-function enhancer that drives higher expression of *KCNB1* in cardiomyocytes. This is the first report of a gain of *de novo* enhancer function causing Mendelian disease. Critically, the tissue-specific gain-of-function regulatory change could be predicted using a deep neural network. Application of a similar framework will enable identification of causal non-coding mutations and affected genes in other rare diseases.

## Introduction

In the last decade, advances in sequencing technologies have produced a wealth of data for understanding the causes of Mendelian disease, however, variant interpretation and identification of disease-causing mutations is still incomplete. Current approaches detect a pathogenic variant in only a third of patients with suspected genetic disease^1–4^, with the vast majority of them located in the coding part of the genome. Candidate gene panels, whole exome sequencing (WES), and certain bioinformatic variant filtering approaches using whole genome sequencing (WGS) data^5^, still frequently miss or disregard non-coding variants. This is partly due to a huge gap in knowledge concerning these parts of the genome. However, just as the majority of common disease variants are thought to affect regulatory, rather than coding sequence^6^, there are likely to be a number of primary disease-causing mutations affecting non-coding regulatory elements that are currently going undetected^7^. Additionally, within efforts to define disease-causing non-coding variants, most emphasis is on detecting variants that disrupt annotated regulatory elements, with few efforts directed at detecting newly created elements and gain-of-function changes. Bridging this gap in knowledge requires better understanding of the mechanisms of non-coding mutations underlying heritable disease, along with improved tools to predict and characterise these changes.

We previously reported a novel inherited cardiac arrhythmia syndrome with widespread ST-segment depression on the electrocardiogram (ECG), termed here ST Depression Syndrome (STDS)^8^. This syndrome is inherited in an autosomal dominant manner and predisposes affected individuals to life-threatening arrhythmias; however, the primary disease-causing mutation remained unknown^9^. Genome-wide association studies (GWAS) for variation in ECG segments, including the PR^10^, QRS^11^, QT^12,13^ and ST^14^ intervals, have all highlighted the role of non-coding variants in the electrophysiology of the heart. Here we report a rare non-coding mutation, shared as a founder allele by affected STDS families, as the underlying genetic cause of this novel cardiac arrhythmia syndrome. By applying a machine-learning, genome-engineering and multi-omics approach developed for the deciphering of GWAS variants^15,16^, we show that the causal mutation, a complex insertion-deletion, repurposes the sequence of a skeletal muscle enhancer to generate a *de novo* gain-of-function cardiomyocyte enhancer that drives increased expression in the heart of the nearby potassium voltage-gated channel encoding gene, *KCNB1*. To our knowledge, this is the first report of a *de novo* enhancer causing Mendelian disease.

## Results

### Clinical findings in families with ST-depression on the ECG

A total of 14 individuals from five UK families (Fig. 1b) were identified as having autosomal dominant ST Depression Syndrome (STDS) (Fig. 1a). Family 1 was described in the original report of this syndrome (Family B in reference 8), with widespread ST-segment depression on the ECG, starting in childhood then stable through life, with incidences of ventricular arrhythmia resulting in cardiac arrest, and atrial fibrillation (AF) and ventricular contractile impairment developing in later life. The proband of Family 2, IV-4, presented with ventricular arrhythmia in mid-adult life and was noted to have widespread ST-segment depression; an implantable cardioverter-defibrillator (ICD) was implanted when she was in her early thirties. In her 50s and 60s she received multiple ICD shocks for symptomatic polymorphic ventricular tachycardia. She died in her early 60s with severe left ventricular (LV) systolic impairment with an ejection fraction of 10%. Two distant relatives, both in their 30s, were found to have the same ECG phenotype: the ECG features were found in IV:1 when he was investigated for chest pain but found to have normal coronary arteries and then in IV:2, who has no symptoms, on family screening. Neither have had significant arrhythmias. Their father (III:1) died in his mid 60s, with ‘abnormal ECG’ noted in clinical records but no other information available. The proband of Family 3, also in his 30s, had the abnormal ECG features detected during an insurance medical, had a structurally normal heart and no arrhythmias. His mother had died suddenly in her early 70s with a history of AF, dilated cardiomyopathy and ST-segment depression reported on her ECG. The proband in Family 4 presented with syncope as a young adult and was found to have the typical ECG changes of widespread, persistent ST-segment depression. Cardiac MRI was normal and no arrhythmia has been documented on an implanted loop recorder. Her mother died suddenly in her early 50s with a clinical record of “sudden cardiac death” but no post-mortem analysis. The proband in Family 5 presented with paroxysmal AF in her 50s. Her sinus rhythm ECGs showed fixed changes typical of familial STDS. Her CT coronary angiogram demonstrated no coronary artery disease, and subsequent cardiac MRI was normal. Her daughter has an identical ECG and is asymptomatic with normal cardiac MRI.

**Fig. 1.**
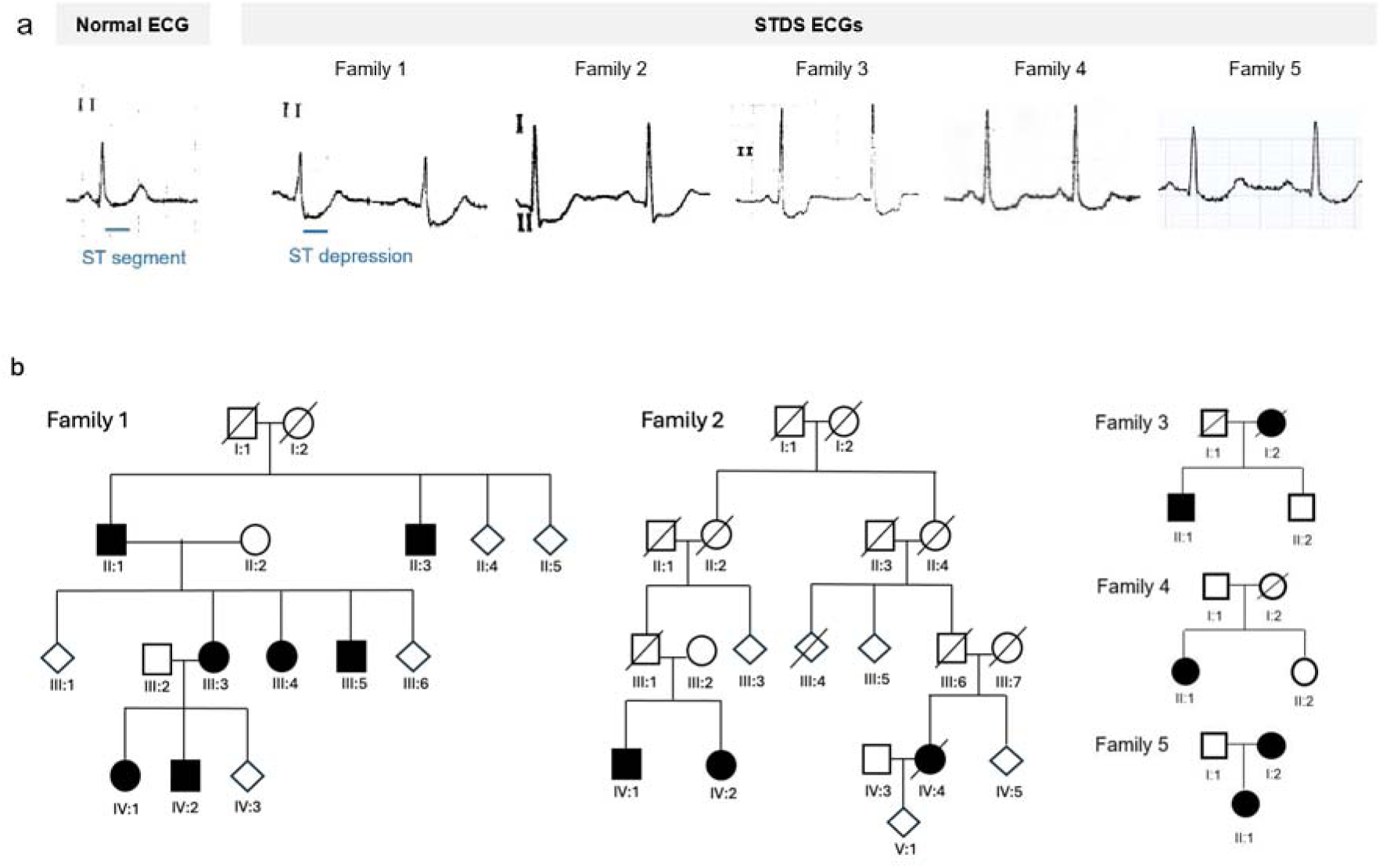
Families with ST-depression syndrome (STDS). **a**, Example ECG lead II traces showing normal versus ST-segment depression in affected members from five independent UK families. **b**, Partial pedigrees of families identified with STDS. Affected individuals are shaded in black. The full pedigrees have not been disclosed to comply with medRxiv’s policies. Full disclosure of the pedigrees is available upon request from the authors.

### Distantly related families with a shared disease locus in a small segment on chromosome 20

To identify regions of interest, independent linkage analyses were performed for each of the two larger families, using data generated from single nucleotide polymorphism (SNP) arrays (Family 1: II:1, II:3, II:5, III:1, III:3, III:4, III:5, III:6, IV:1 and IV:2, Family 2: III:5, IV:4, IV:5 and V:1) and WGS (Family 2: IV:1). For Family 1, two regions of the genome on chromosomes 12 and 20 showed complete linkage (LOD = 2.41) with ST-segment depression (Fig. 2a, Supplementary Table 1); all other chromosomal regions were excluded with LOD < −2.0. Family 2 did not share the linkage region on chromosome 12, but an overlapping region showing perfect segregation (hg38, chr20:48,212,554-50,048,337, LOD = 1.52) was detected on chromosome 20 (Fig. 2a, Supplementary Table 1), indicating that these two families likely share a single causative locus.

**Fig. 2.**
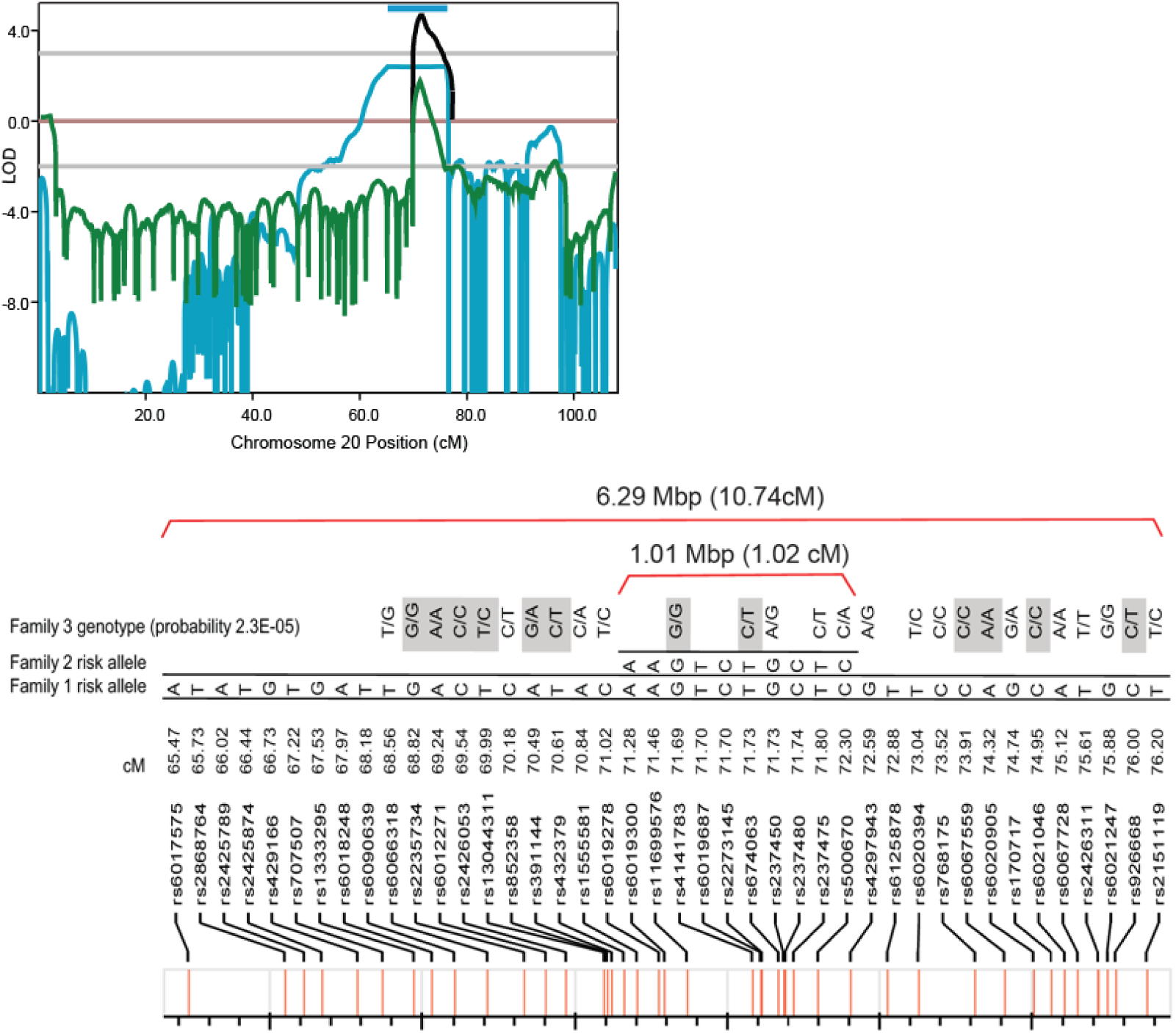
A shared, identical-by-descent, disease locus on chromosome 20. **a**, Linkage analysis identified a single linkage region shared by family 1 (blue) and family 2 (green) on chromosome 20. The blue bar denotes the 6.29Mbp region in complete linkage in Family 1 and is displayed in b). **b**) 3-4 markers were selected per cM and the linkage boundaries were determined based on recombination events. The risk haplotypes of families 1 & 2 are shown and are seen to be identical in a ∼1Mbp region; as the haplotype is rare this indicates identity-by-descent, allowing combining of the LOD scores (black). Genotypes of the affected family 3 member are shown above; SNPs in linkage with each other are highlighted in grey and excluded from probability estimation for Family 3.

To explore the possibility that these two STDS families could be distantly related we next examined informative SNPs from the array genotyping to define the haplotypes segregating with disease in the regions of complete linkage in each of Family 1 and 2. In Family 1, this revealed a 6.29 Mbp (10.74cM) segment carried by each of the 7 affected members and not carried by the unaffected family members (Fig. 2b, Supplementary Fig. 1a,b). In Family 2, haplotype analysis revealed a 1.01 Mbp (1.02 cM) region similarly showing complete linkage with STDS, delimited by recombination events within the family (Fig. 2b, Supplementary Fig. 1c,d). The haplotypes in this shared region of complete linkage were identical in Family 1 and Family 2.–Inspection of this haplotype shows that it is rare, carried by 0.006% of European ancestry individuals in UKBiobank (Supplementary Fig. 2). Thus, this finding provides robust evidence that this sharing has not arisen by chance, but rather Family 1 and 2 are distantly related, i.e. share a common ancestor, such that this shared region is identical-by-descent (IBD). Confirmation of a shared IBD region allows linkage data from the two families to be combined for estimation of minimal LOD score; this gives a combined LOD of 4.26 (Fig. 2a).

In Family 3, whole genome sequence data was available on the proband, but not SNP array data. Genotypes were therefore extracted for informative SNPs in the linkage region on chromosome 20. This revealed an ∼ 4.3 Mbp segment in which the disease-associated allele in the risk haplotype in Family 1 was always present in the Family 3 proband’s genotype. To estimate the probability that this might have arisen by chance, we examined SNPs not in LD with one another across this region and calculated the combined likelihood of carriage of at least one copy of the relevant allele. This showed that the probability that this might have arisen by chance in less than 2.3 x 10^-05^, again indicating that this segment has been passed down IBD from a common ancestor. Taken together these analyses indicate that the disease locus in these families lies on chromosome 20 within the small ∼1Mbp region defined by the extent of the IBD segment and haplotype shared by all three.

### Affected family members share a single non-coding variant

Variant calling and quality-based filtering with Platypus^17^, using whole genome sequence data for the six affected individuals from families 1, 2 and 3, resulted in an aggregated dataset containing 6,926,029 variants (6,133,943 SNPs and 792,086 indels). Of these variants, there were 22 novel heterozygous variants shared by the 6 selected affected individuals that were absent from both the general population (gnomAD and 1000G) and the control cohort (Supplementary Table 2). None of these variants affected coding sequence and only one was in the region defined by genetic linkage and the extended area of IBD and haplotype sharing. This variant was a single, small, complex deletion and insertion located in the ∼1Mbp shared region on chromosome 20 and carried on the shared affected haplotype (see Supplementary Table 3): NC_000020.11: g.49356862-49356878delinsTCCC, i.e. comprising loss of 17 residues and gain of four others, referred to hereafter as delinsTCCC (Fig. 3). The delinsTCCC variant was absent in all databases searched, including gnomAD and the 100KGP project.

**Fig. 3.**
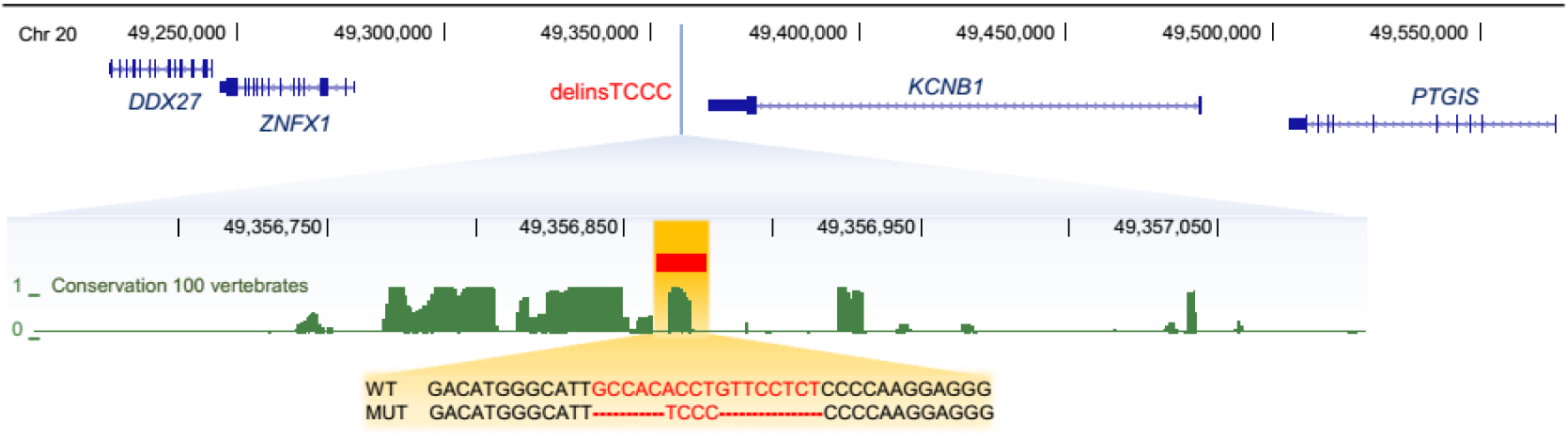
The delinsTCCC variant and flanking sequence. Sequence and relative chromosome 20 position (hg38) of the indel variant. Green conservation peaks represent PhastCons conservation predictions using 100 vertebrates.

Sanger sequencing was used to confirm that this variant was absent in unaffected relatives (n=4, Family 1: III:6, Family 2: III:5, IV:5 and V:1) and present in all affected cases from families 1, 2 and 3 (n=12, Family 1: II:1, II:3, III:3, III:4, III:5, IV:1 and IV:2, Family 2: IV:1, IV:2 and IV:4, Family 3: II:1). Finally, Sanger sequencing revealed that the same delinsTCCC was present in both probands from the two additional families subsequently ascertained, Families 4 & 5.

### In silico analysis of the delinsTCCC variant

The delinsTCCC variant lies in a highly conserved intergenic region of the genome, 6,999 bp downstream of the nearest annotated gene, the potassium channel encoding gene *KCNB1* (Fig. 3) and 78.8 kbp upstream of the nearest transcription start site (*ZNFX1*). Given the non-coding location of this variant we reasoned this region could play a role in transcriptional regulation of genes influencing the ST-segment. Consistent with this hypothesis, it is positioned within 50 kbp of two independent sentinel SNPs identified by a GWAS for ST-segment amplitude^14^ (Fig. 4 and Supplementary Dataset 1). Application of a platform designed to decode GWAS signals^15,16^ by considering regulatory, splicing and coding mechanisms, attributed these signals to two moderately linked candidate causal variants (rs6012624, rs6019764, r^2^: 0.715 [EUR]), which both lie in open chromatin in cardiomyocytes. Using a deep convolutional neural network (CNN)^15,18^ trained to predict cell type-specific chromatin activity from sequence we predicted that both GWAS variants alter chromatin accessibility in cardiomyocytes (Supplementary Dataset 1). Published low-resolution (20 kbp) chromosome conformation capture (3C) Hi-C data from cardiomyocytes differentiated from induced pluripotent stem cells (iPSC) and embryonic stem cells (ESC)^19^ shows that one of the predicted causal GWAS variants and the delinsTCCC variant lie in a region of chromatin that interacts with the *KCNB1* promoter, but not the *ZNFX1* promoter (Fig. 4a). These results suggest that altered gene regulation at this locus can affect the ST-segment and may be responsible for STDS.

**Fig. 4.**
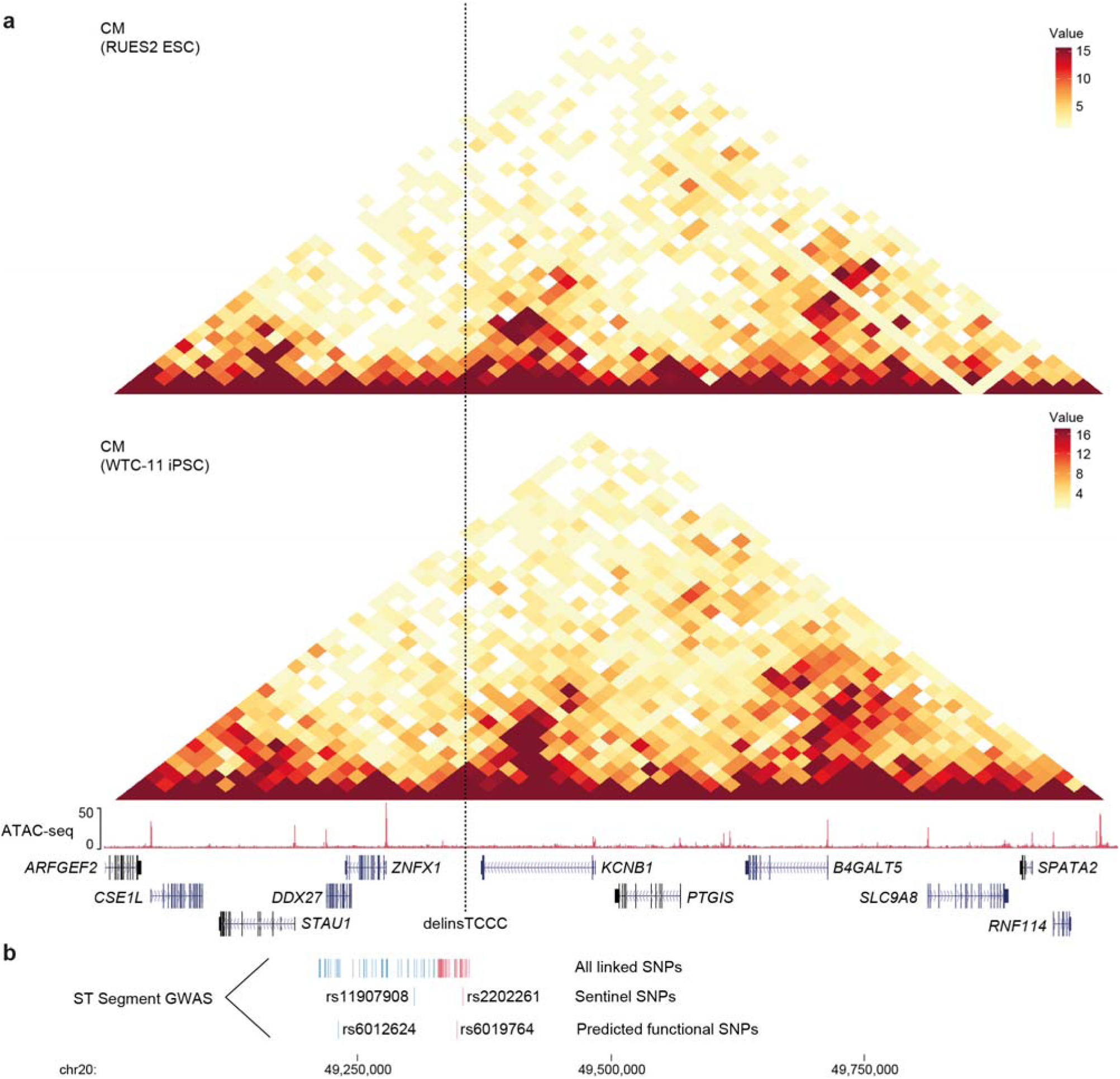
*In silico* analysis of the novel delinsTCCC variant and flanking sequence. **a**, HiC interaction plots for ESC and iPSC derived cardiomyocytes, ATAC-seq peaks for iPSC derived cardiomyocytes, genes and delinsTCCC location. **b**, SNPs in linkage (r^2^ ≥0.8 [EUR]) with ST-segment GWAS sentinel SNPs rs11907908 (blue) and rs2202261 (red).

Given the sequence conservation surrounding the delinsTCCC variant and the nearby predicted regulatory GWAS SNPs, we investigated Assay for Transposase-Accessible Chromatin-sequencing (ATAC-seq) data from ESC derived cardiomyocyte progenitors^19^ and iPSC derived cardiomyocytes from unaffected individuals^20^, and single cell ATAC-seq (scATAC-seq) from developing embryonic heart^21^ for the presence of open chromatin, which is associated with enhancers. This analysis revealed that the delinsTCCC site does not lie in a region of open chromatin in cardiac cells (Fig. 4a and Fig. 5a) with the nearest open chromatin located ∼12 kbp upstream; given the Hi-C chromatin structure we named this site E-139 as it is a putative enhancer and 139 kbp from the *KCNB1* promoter (Fig. 5a, Supplementary Fig. 3). A broader survey of 95 ENCODE DNase1-seq open chromatin datasets^22,23^ and 126 developmental scATAC-seq datasets^21^ shows this region is accessible in skeletal muscle cells and multiple carcinomas (Supplementary Fig. 4) and is designated as a distal enhancer like sequence by ENCODE. Recent work has shown how sequence changes in enhancers can lead to loss of tissue-specificity by optimizing transcription factor binding affinity; including for cardiac enhancers that regulate ECG traits^24,25^. Given the chromatin accessibility in non-cardiac muscle cells and the nearby ST-segment GWAS signal we hypothesized delinsTCCC may lead to a change in enhancer tissue-specificity, to generate a cryptic *de novo* cardiomyocyte enhancer. Supportive of this hypothesis, the predictions from the deep CNN found that delinsTCCC had the potential to increase chromatin accessibility in cardiomyocytes and smooth muscle cells but not in carcinomas (Fig. 5b, Supplementary Fig. 5). *In silico* mutagenesis of each base pair across the delinsTCCC region showed that a TCCC motif (Fig. 5c), corresponding to several transcription factor families, was important for the predicted gain in accessibility. Consistent with binding-affinity optimization, both ETS and NFKB showed stronger motif matches to the variant sequence than to reference sequence; whereas the E2F motif is only found in the variant sequence (Fig. 5d). These findings strengthen the evidence for this disease-causing variant while supporting the hypothesis for a *de novo* cardiomyocyte enhancer.

**Fig. 5.**
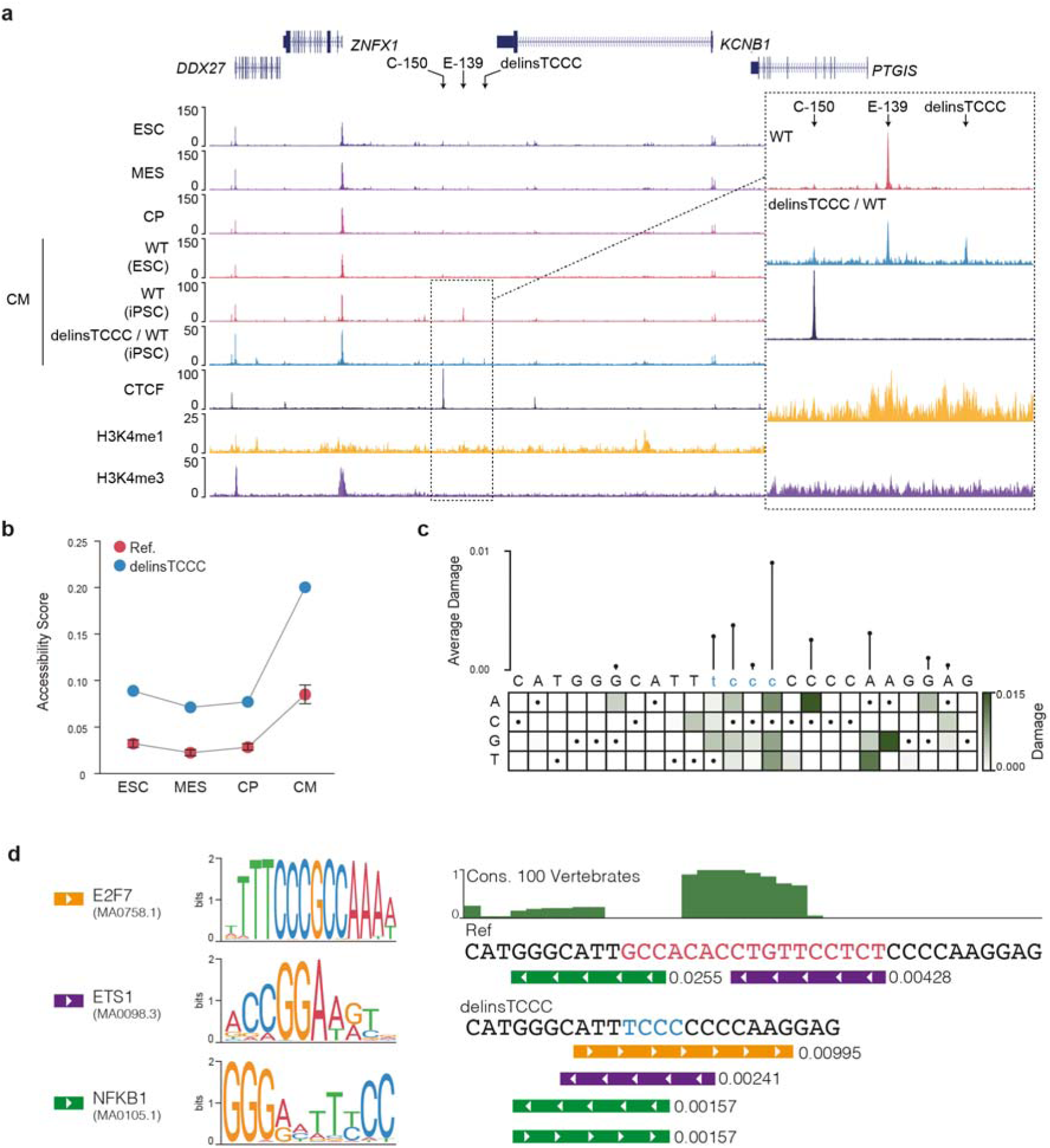
Accessibility analysis of the delinsTCCC variant sequence. **a**, ATAC sequencing analysis of published data (GSE89895, GSE106690) for cardiomyocytes (CM) and progenitors, including mesoderm (MES) and cardiac progenitors (CP), derived from differentiation of embryonic stem cells (ESC), or induced pluripotent stem cells (iPSC). Cardiomyocytes either lacked (WT) or were heterozygous (delinsTCCC/WT) for the delinsTCCC variant. CTCF ChIP-seq (Erythroid, GSE125926) shows tissue-invariant boundary elements. H3K4me1 and H4K4me3 CUT&RUN in delinsTCCC heterozygous cardiomyocytes show enhancers and promoters respectively. Inset region is chr20:49,327,000-49,367,000 (hg38) with 5-pixel window. **b**, deep CNN accessibility predictions. Error bars denote one standard deviation of accessibility score for the 13 possible 1 kbp sequences covering the indel site. **c**, *in silico* mutagenesis for the delinsTCCC site indicating a possible motif including the TCCC insert. **d**, left panel shows the colour coded FIMO derived predicted motifs from both the reference and variant delinsTCCC sequence. The right panel shows the PhyloP sequence conservation plotted over the reference sequence (Ref) with the deleted sequence high-lighted in red and with the position of motif matches and FIMO p-values below. The delinsTCCC variant sequence is shown below with the 4 base-pair insertion highted in blue and with the position of motif matches and FIMO p-values below, showing the creation of a strong E2F7 motif and the rearrangement and increase in scores for pre-existing motifs.

### delinsTCCC generates a cardiac enhancer

To test if delinsTCCC generates an open chromatin site in human cardiomyocytes, we generated iPSC lines (Supplementary Fig. 6) heterozygous for the delinsTCCC variant using CRISPR-Cas9 genome editing (Supplementary Fig. 7). ATAC-seq in cardiomyocytes differentiated from these iPSC showed the presence of a *de novo* open chromatin element which contained the delinsTCCC variant (Fig. 5a). Allelic analysis of reads in this region indicated that over 84% of the reads came from the mutated allele, implying that the peak was specific to the delinsTCCC sequence (Supplementary Fig. 7). Relative levels of histone H3 lysine-4 mono- and tri-methylation (H3K4me1, H3K4me3) can be used to distinguish enhancers and promoters^26^. We performed Cleavage Under Targets and Release Using Nuclease (CUT&RUN)^27^ for both H3K4me1, which is primarily at enhancers, and H3K4me3, which is primarily at promoters, in the heterozygous mutant cardiomyocyte cells. Higher levels of H3K4me1 relative to H3K4me3 were detected at the *de novo* element and E-139 (Fig. 5a, Supplementary Fig. 8), consistent with ENCODE ChIP-seq for these marks in smooth muscle cells (Supplementary Fig. 4), indicating that they are both likely to be enhancers.

To determine if the delinsTCCC sequence and the E-139 element function as enhancers we placed each, as well as the equivalent wild-type sequence for the delinsTCCC site (WT), separately upstream of the silent E1b minimal promoter and a GFP reporter gene and generated Tol2-induced transgenic zebrafish embryos. Both the *tg(E-139:GFP)* and the *tg(delinsTCCC:GFP)* transgenic fish showed strong GFP expression in the heart, spread throughout the ventricles in a fairly consistent manner (Fig. 6a,b, Supplementary Table 4). By comparison, the *tg(WT:GFP)* transgenic fish displayed low or negligible GFP expression in this region. Expression in certain other parts of the fish (e.g. the eye) was in keeping with commonly observed autofluorescence (Supplementary Fig. 9).

**Fig. 6.**
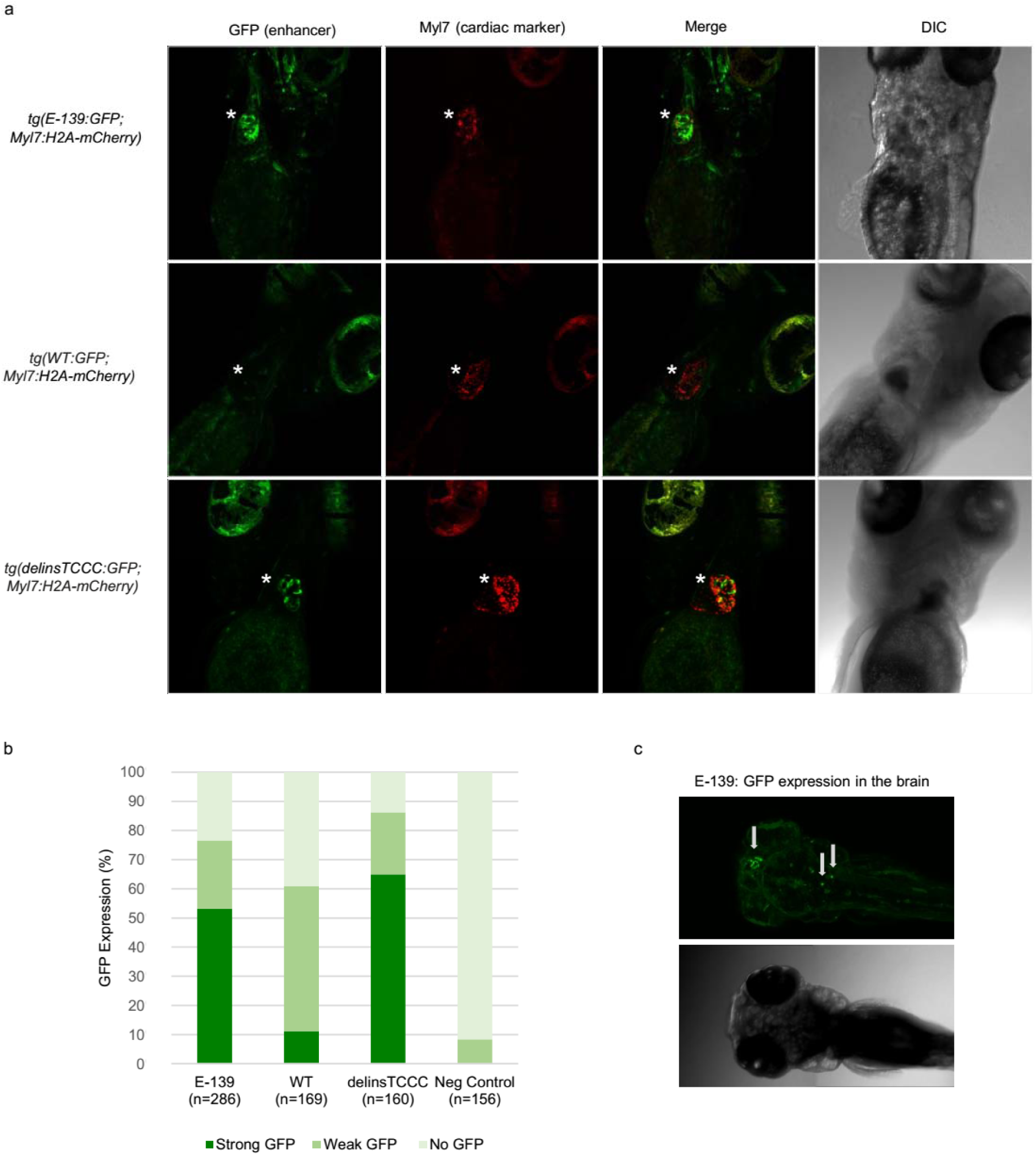
Enhancer variant expression in zebrafish embryos. **a**, GFP expression for transgenic fish containing the E-139, wild type (WT) or delinsTCCC variant sequence. The asterisk indicates the location of the heart. The Myl7 cardiac marker is overlaid in red. DIC = differential interference contrast. **b**, Total GFP levels observed in the hearts of transgenic zebrafish; n represents the total number of transgenic zebrafish embryos per category. **c,** E-139 transgenic fish displayed strong expression in the brain indicated by the arrows.

In addition to the heart, the *tg(E-139:GFP)* fish also displayed strong expression in other parts of the fish including the head (Fig. 6c, Supplementary Fig. 10). Consistently, E-139 was accessible in numerous cerebrum cell-types in ATAC-seq from developing human brains^21^ (Supplementary Fig. 3). This is interesting as the closest gene, *KCNB1,* is known to play a significant role in the brain^28^. Therefore, while the E-139 element appears to be involved in more ubiquitous expression, the delinsTCCC variant seems to specifically drive gene expression in the heart.

### delinsTCCC drives higher *KCNB1* expression in cardiomyocytes

To identify the target gene(s) of the *de novo* delinsTCCC enhancer, we generated high-resolution 3C interaction profiles for the three closest active genes (*KCNB1*, *ZNFX1*, *PTGIS*) with nuclear-titrated Capture-C^29^ using wild-type and heterozygous delinsTCCC iPSC derived cardiomyocytes. Capture-C from the *KCNB1* promoter in the wild-type cells showed strong interaction with E-139 and a proximal CTCF site (C-150) situated 150 kbp upstream of the promoter (Fig. 7). Neither *ZNFX1* nor *PTGIS* showed strong interaction with this region (Supplementary Fig. 11). Capture-C from *KCNB1* in heterozygous delinsTCCC cells, showed significantly higher levels of interaction with fragments covering E-139 compared to the wild-type cells (Fig. 7; two-sided Wilcoxon matched-pairs signed rank test, p<0.0001). To exclude the possibility that these elements interact with more distal genes, we next performed Capture-C experiments from both the delinsTCCC enhancer and E-139. The delinsTCCC site did not show a strong interaction with the *KCNB1* promoter, nor any other promoter, in either wild-type or heterozygous cells. The E-139 enhancer interacted specifically with the *KCNB1* promoter and the level of interaction was significantly increased in the presence of the delinsTCCC allele (two-sided Wilcoxon matched-pairs signed rank test, p=0.0094).

**Fig. 7.**
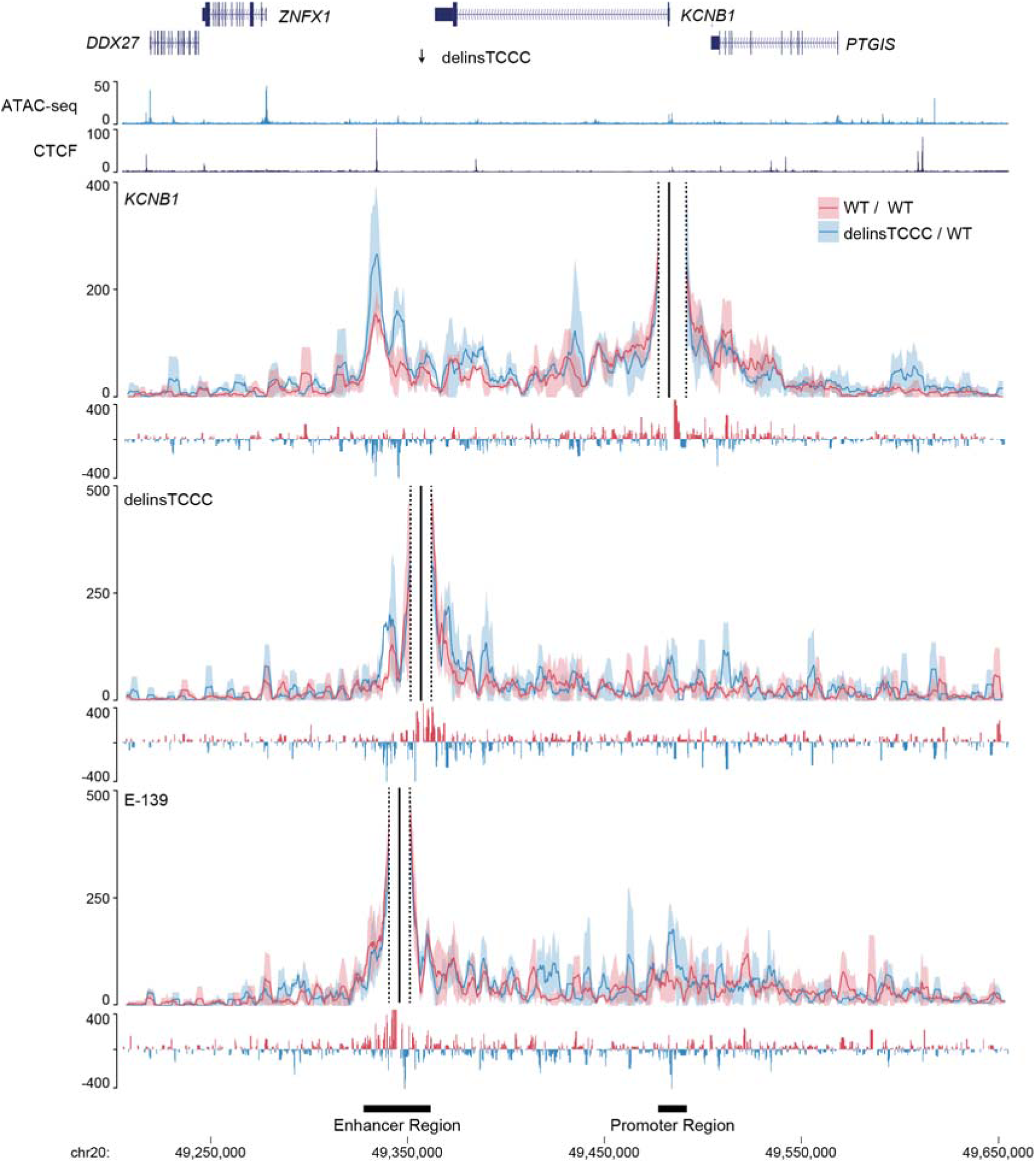
Chromatin conformation and interaction assays. ATAC sequencing analysis of experimental data from iPSC derived cardiomyocytes (CM) heterozygous (delinsTCCC/WT) for the delinsTCCC variant. CTCF ChIP-seq shows boundary elements. Capture-C from the *KCNB1* promoter, delinsTCCC enhancer, and the E-139 enhancer. Solid lines show mean (n=3 independent samples) with one standard deviation (shading). Subtraction tracks show per *Dpn*II fragment difference. Enhancer and promoter regions indicated below display regions of significant difference.

Finally, to determine whether the increased E-139/*KCNB1* interaction in the presence of the delinsTCCC variant led to higher gene expression, we performed quantitative real-time PCR experiments using iPSC derived cardiomyocytes. Three independent heterozygous clones (C6, C68 and C93), each with multiple differentiations, collectively showed markedly increased *KCNB1* expression (two-sided Mann-Whitney test, p=0.0015) (Fig. 8a).

**Fig. 8.**
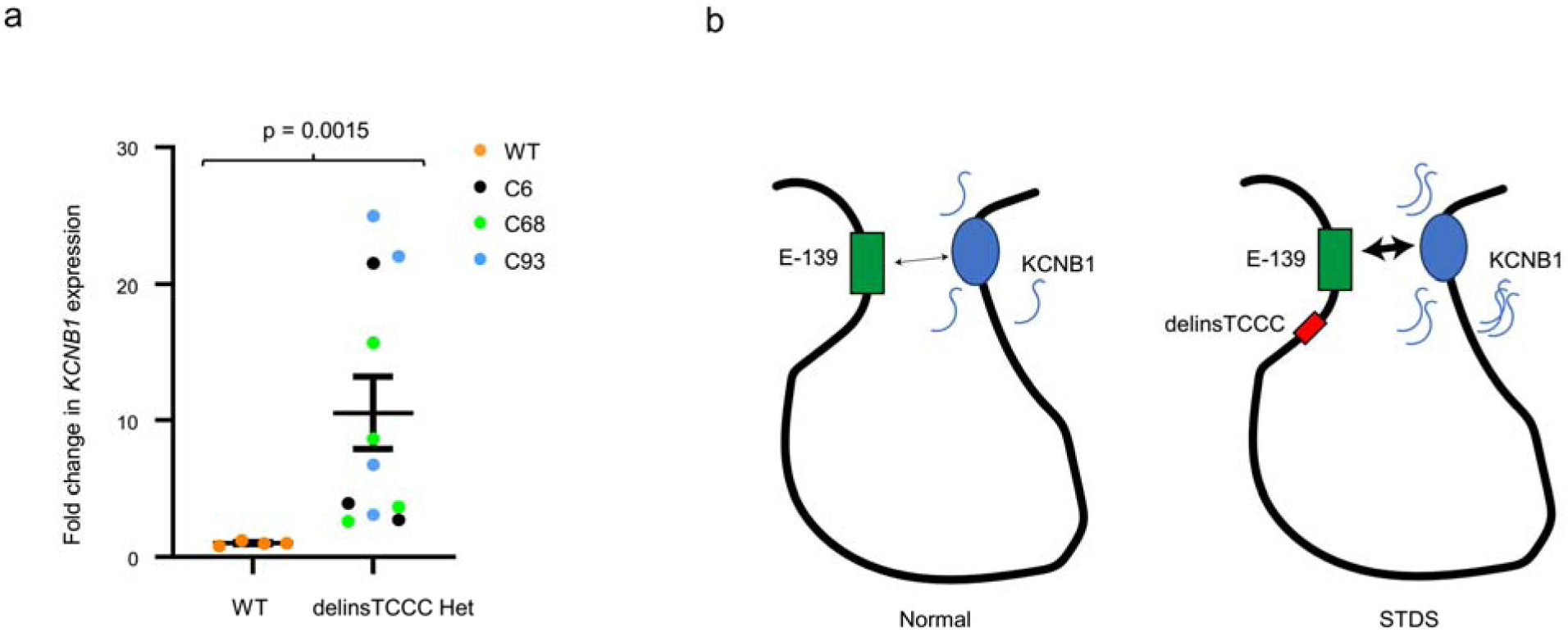
KCNB1 upregulation by the delinsTCCC variant. **a,** Fold change in *KCNB1* expression versus the housekeeping gene (*GAPDH*) detected by qPCR for reference (WT n=4 independent samples/differentiations) and heterozygous delinsTCCC clones (C6 n=3, C68 n=4, C93 n=4 independent samples/differentiations). The mean for each group is represented by the black horizontal line, bars show standard error of the mean and the p-value from two-sided Mann-Whitney test is indicated. **b,** Model illustrating a proposed regulatory mechanism with the delinsTCCC variant indirectly enhancing *KCNB1* transcription.

To investigate differences in allelic expression, we identified a SNP (rs2229006) within the coding sequence of *KCNB1* (exon 2, NM_004975.4) that was heterozygous in the parental iPSC cell line. By combining public phase data and Nanopore sequencing we determined that the variant allele, rs2229006-C, was in *cis* with the edited delinsTCCC allele in clone C6 (Supplementary Fig. 12). RNA sequencing showed a bias for *KCNB1* transcripts arising from the chromosome containing the delinsTCCC variant, with the rs2229006-C allele being 3.8-fold more abundant than the reference rs2229006-G allele (Supplementary Table 5). These findings corroborate a mechanism whereby a *de novo* enhancer, created by the delinsTCCC variant, leads to cardiac specific *KCNB1* upregulation through an increased interaction between E-139 and the *KCNB1* promoter (Fig. 8b).

## Discussion

We have utilised a multi-omics, genome engineering and machine learning^15,16^ approach to identify and characterise a novel gain-of-function non-coding disease-causing variant responsible for a recently described cardiac arrhythmia syndrome, STSD^8,30^. This syndrome is associated with widespread ST-segment depression on the ECG and predisposes affected individuals to cardiac events including sudden cardiac death and heart failure. The responsible variant, delinsTCCC, generates a cryptic cardiomyocyte enhancer which drives higher expression of the potassium channel encoding gene, *KCNB1*.

Cardiac arrhythmia syndromes are most commonly associated with variants in ion channel genes^31^. Consistent with this, the causal variant identified in this study enhances the expression in the heart of the *KCNB1-*encoded, pore-forming subunit of the voltage-gated potassium channel Kv2.1. This channel acts as a delayed rectifier, propagating current in a wide range of electrically active cell types across many organ systems. Kv2.1 plays an important role in the repolarisation phase of the action potential in rodent hearts^32,33^, is expressed in human atrial^34–36^ and ventricular cells^35,36^, and binds ion channel beta-subunits from the KCNE family^37^ known to contribute to other cardiac arrhythmia syndromes such as the Long-QT syndrome^38,39^. However, rather than causing cardiac disease, pathogenic *KCNB1* coding variants have been shown to cause neurological disorders, specifically forms of developmental and epileptic encephalopathy^40^. Much attention has been devoted to the study of Kv2.1 in the human brain, largely due to its high expression in this region^41^.

Interestingly, our study identified the regulatory element proximal to the delinsTCCC enhancer, E-139, as an interactor of the *KCNB1* gene promoter and we confirm its ability of driving reporter gene expression in zebrafish brain. The broad clinical spectrum of subjects with different *KCNB1* variants emphasises the cell-type specific impact of regulatory changes which determine which organs are affected, and hence the phenotypes of these inherited conditions.

The hallmark phenotype of resting ST-segment depression on the surface ECG in STDS implies transmural heterogeneity of currents underlying the cardiac action potential, thereby creating a transmural gradient (analogous to the ST-segment shifts seen acutely in the context of subendocardial ischaemia or myocardial infarction)^42,43^. Therefore, we hypothesise that the delinsTCCC enhancer results in differential increases in KCNB1 expression, and hence Kv2.1 activity, in different layers of the myocardium – a property commonly associated with cardiac ion channels^44^. A shorter endocardial action potential, induced by an endocardial increase in expression of the Kv2.1 channel, would promote current flow from epicardial sites with longer action potentials to generate ST depression on the surface ECG. This is the inverse of the gradient seen in Brugada Syndrome which promotes localised ST elevation associated with different transmural action potential durations where the epicardial action potential duration is relatively short ^45,46^. These gradients require electrotonic uncoupling to be maintained. Indeed, the fact that the ST depression develops 80ms after the J point would be compatible with such differences developing in phase 3 and 4 of the action potential when Kv2.1 is active. Such gradients set up potential sites for phase 2 re-entry following an ectopic beat initiating the polymorphic ventricular arrhythmias described in this rare familial condition^47^.

The delinsTCCC variant underlying STDS adds to the growing knowledge of non-coding regulatory variants associated with ECG traits. Previous reports, including a number of GWAS^10–14^, have identified non-coding regions important for the ST^14^, QT^12,13,48–50^, PR^10,51–53^ and QRS^11,51–53^ intervals. However, identifying specific target genes and their associated functional effects is still a huge challenge. This has limited the detection and inclusion of pathogenic non-coding variants in clinical practice. Furthermore, while pairing genes to their regulatory elements can be carried out using a combination of chromosome conformation capture techniques^54^ and open chromatin atlases^21^, these methods are of little use for the identification of *de novo* enhancer variants. Therefore, our study highlights the value of combining these rich resources with ongoing ‘classic’ family studies, cellular models and machine learning to improve non-coding variant detection and validation.

Gain-of-function mechanisms are an important consideration when trying to decipher non-coding effects. Despite reports of gain-of-function variants within existing enhancers in common disease, rare disease and malignancy^55–57^, we believe this is the first description of an entirely *de novo* cryptic enhancer causing a Mendelian disorder. Variants creating *de novo* regulatory elements may conceivably be more common than expected. Recently, a single nucleotide variant which causes a-thalassemia was fully characterised as generating a gain-of-function cryptic promoter which blocks interaction between the two a-globin encoding genes and their cognate enhancers^58^. Additionally, in T-cell acute lymphoblastic leukaemia, gain-of-function deletions upstream of the *TAL1* oncogene generate a *de novo* enhancer which drives higher gene expression^59^. Similarly, in several autosomal dominant heritable diseases, including brachydactyly-anonychia^60^, Keratolytic Winter Erythema^61^, Haas-type polysyndactyly and Laurin-Sandrow syndrome^62^, as well as syndactyly and craniosynostosis^63^, duplications of developmental gene enhancers cause disease through gain-of-function.

Intriguingly, our 3C approach did not detect significant changes in the interaction between the *de novo* cryptic enhancer and the *KCNB1* promoter. This suggests that the delinsTCCC enhancer may function in an indirect manner, by promoting stronger interaction between E-139 and the *KCNB1* promoter. Recent work to dissect the role of the five independent elements of the mouse alpha-globin super enhancer has shown that while some elements look like classical tissue-specific enhancers in reporter assays, they instead act as facilitators for nearby enhancers in their native context^64^. This is consistent with the tissue-specific GFP expression driven by the delinsTCCC enhancer in the Zebrafish reporter assay. This mechanism is compatible with two current models for gene regulation. In the first model, the loop extrusion model^65^, the delinsTCCC enhancer could be recruiting additional cohesin^66^, promoting an increase in the frequency of the interaction between *KCNB1* and E-139. In contrast to this model, the second involves an increase in bound transcription factors leading to more frequent or stable phase-separation^67^. Further research is required to better understand these models and their contribution to disease.

In addition to specific cardiovascular conditions, including cardiac arrythmias^8^, ischemia or coronary heart disease^68^, ST-segment depression on the ECG is associated with an increased risk of unexplained sudden cardiac death in the general population^69^. GWAS have identified numerous common variants that are associated with ST-segment variability^14^, some of which are near the delinsTCCC variant. Therefore, additional studies of non-coding variants associated with ECG alterations from both GWAS and Mendelian disease will likely improve our general understanding of both gene regulation and cardiovascular disease.

## Data Availability

All data produced in the present work are contained in the manuscript

## Acknowledgements

The authors are grateful to Nadav Ahituv from the University of California, San Francisco for gifting the E1b-GFP-Tol2 vector. We would also like to thank Charlotte Ives, Inherited Arrhythmia Clinical Nurse Specialist, Barts Health NHS Trust, London, and Ellie Quinn from Royal Brompton and Harefield Hospitals for assistance with patient records. Additionally, we would like to thank the Oxford Genomics Centre at the Centre for Human Genetics for the generation and initial processing of sequencing data.

This work was supported by funding from the British Heart Foundation (BHF), Wellcome Trust (nos.090532/Z/09/Z and 203141/Z/16/Z to H.W.) and the National Institute for Health Research (NIHR) Oxford Biomedical Research Centre. The views expressed are those of the authors and not necessarily those of the NHS, the NIHR or the Department of Health or Wellcome Trust. J.R.H received funding from a Wellcome Strategic Award (no. 106130/Z/14/Z), Wellcome Discovery Award (225220/Z/22/Z) and Medical Research Council Core Funding (no. MC_UU_00016/14 and MC_UU_00029/3). R.S. was supported by a Wellcome Doctoral Programme (no. 203728/Z/16/Z). R.A.B was funded by a Sir Henry Wellcome Fellowship (no. 209181/Z/17/Z). S.D. and S.N. were supported by a BHF Fellowship (no. FS/1735/32929), MRC project grant (no. MR/S01019X/1) and by Leducq Foundation grant 18CVD03. P.L. was supported by UCL/UCLH and Barts NIHR Biomedical Research Centres. H.W.’s laboratory is supported by the British Heart Foundation’s Big Beat Challenge award to CureHeart (BBC/F/21/220106).

## Author contributions

C.P.D.V, D.J.D., A.J.S., S.N., P.R., M.E.G., R.A.B., M.L., C.W., A.T. performed experiments.

C.P.D.V, D.J.D., A.T.P., A.J.S., S.N., E.G., A.G., M.F., R.S., D.P., S.L, S.S, H.R. processed data and performed analyses.

C.P.D.V, D.J.D. designed experiments.

C.P.D.V., D.J.D., A.T.P., A.J.S., S.N, E.G., A.G., M.F., P.R, D.P. wrote the manuscript and made figures.

E.O., R.H., E.R., G.S, P.L., H.W. collected and evaluated clinical data. C.P.D.V, A.J.S. cultured iPSC derived cardiomyocytes.

S.N. performed zebrafish investigations.

P.R. produced CRISPR-Cas9 edited iPSC cell lines.

M.L, C.W., A.T. performed nanopore sequencing.

M.F., B.D., D.B., J.C.T., C.R., S.D.V., J.R.H., H.W. provided supervision.

H.W., J.R.H., acquired funding, oversaw the work, and revised the manuscript.

H.W. conceived the work.

## WGS500 Consortium membership: names and affiliations of authors

Steering Committee: Peter Donnelly (Chair)^1^, John Bell^2^, David Bentley^3^, Gil McVean^1^, Peter Ratcliffe^1^, Jenny C. Taylor^1,4^, Andrew Wilkie^4,5^

Operations Committee: Peter Donnelly (Chair)^1^, John Broxholme^1^, David Buck^1^, Jean-Baptiste Cazier^1^, Richard Cornall^1^, Lorna Gregory^1^, Julian Knight^1^, Gerton Lunter^1^, Gil McVean^1^, Jenny C. Taylor^1,4^, Ian Tomlinson^1,4^, Andrew Wilkie^4,5^

Sequencing & Experimental Follow up: David Buck (Lead)^1^, Christopher Allan^1^, Moustafa Attar^1^, Angie Green^1^, Lorna Gregory^1^, Sean Humphray^3^, Zoya Kingsbury^3^, Sarah Lamble^1^, Lorne Lonie^1^, Alistair T. Pagnamenta^1^, Paolo Piazza^1^, Guadelupe Polanco^1^, Amy Trebes^1^ Data Analysis: Gil McVean^1^ (Lead), Peter Donnelly^1^, Jean-Baptiste Cazier^1^, John Broxholme^1^, Richard Copley^1^, Simon Fiddy^1^, Russell Grocock^3^, Edouard Hatton^1^, Chris Holmes^1^, Linda Hughes^1^, Peter Humburg^1^, Alexander Kanapin^1^, Stefano Lise^1^, Gerton Lunter^1^, Hilary C. Martin^1^, Lisa Murray^3^, Davis McCarthy^1^, Andy Rimmer^1^, Natasha Sahgal^1^, Ben Wright^1^, Chris Yau^6^

^1^The Centre for Human Genetics, Roosevelt Drive, Oxford, OX3 7BN, UK

^2^Office of the Regius Professor of Medicine, Richard Doll Building, Roosevelt Drive, Oxford, OX3 7LF, UK

^3^Illumina Cambridge Ltd., Chesterford Research Park, Little Chesterford, Essex, CB10 1XL, UK

^4^NIHR Oxford Biomedical Research Centre, Oxford, UK

^5^Weatherall Institute of Molecular Medicine, John Radcliffe Hospital, Headington, Oxford OX3 9DS, UK

^6^Imperial College London, South Kensington Campus, London, SW7 2AZ, UK

