## Supplementary material for "An autosomal dominant cardiac arrhythmia syndrome, ST Depression Syndrome, is caused by the *de novo* creation of a cardiomyocyte enhancer": Methods

**ST-segment depression syndrome families**

This study includes four UK families with widespread ST-segment depression on the ECG. The phenotype of family 1 has previously been described^1^. Through further clinical screening four additional families with the same clinical features were identified. All individuals participating in this study provided informed consent. Research ethics approval was given by the Oxfordshire Research Ethics Committee B: titled “Molecular Genetic Studies of Individuals and Families at risk of Inherited Cardiac Disease,” approval reference 09/H0605/3. DNA was extracted from peripheral blood or saliva.

**DNA sequencing**

Whole genome sequencing for three affected individuals was performed at the Wellcome Centre for Human Genetics. Two of those, from family 1 (Fig 1b II:5 and III:3), were sequenced in 2011 on a HiSeq2000 instrument as part of the WGS500 study^2^. Base-calling was performed with Bustard v1.9.3. The third, from family 3 (Fig 1b II:1), was sequenced with a NovaSeq 6000 system using Illumina bcl2fastq v2.20.0 for base-calling and bwa mem v0.7.15 for mapping to the human genome assembly GRCh38 (hg38). Two additional genomes from affected members in family 1 (Fig. 1b II:1 and IV:1) and one from family 2 (Fig. 1b IV:1) were sequenced by Edinburgh Genomics using the HiSeq X machine and the TruSeq PCR-free protocol. BCBio-Nextgen (0.9.7) was used for base calling and aligning to hg38.

Sanger sequencing was used to confirm the presence or absence of the delinsTCCC variant in additional family members from families 1-3, and the proband from Family 4.

Nanopore sequencing was performed at the Wellcome Centre for Human Genetics for PCR amplicons amplified with the KAPA Long Range PCR system (KK3502). For sequencing, purified amplicons were normalised to 200-250 ng in 24 µl nuclease free water and processed following the Native Barcoding Genomic DNA protocol (catalogue numbers SQK-LSK109 and EXP-NBD104; revision NBE_9065_v109_revV_14Aug2019) from Oxford Nanopore Technologies (England, UK). Each sample was ligated with a unique native barcode prior to equimolar pooling. The multiplexed pool was sequenced on a Flongle flowcell for 24 hours. Base-calling and demultiplexing were performed using Guppy (version 4.2.2) under GNU Parallel^3^, discarding low quality reads and reads which included mid strand barcodes. The reads for each sample were then mapped to hg38 using the version of minimap2 bundled with Guppy. Samtools 1.4.1 was used to merge and sort to produce a single BAM per sample.

**SNP array data acquisition**

Genotyping of samples was performed with Illumina HumanCytoSNP-12v2 BeadChips for affected (n=6) and unaffected (n=4) individuals and called with Beadstudio using a human genome build 38 map files. Call rates were over 98.5 % for 13/14 samples. One DNA sample obtained from saliva, from family 1 member IV1, had a call rate of 97.3 % and was known to have a low A260/230 ratio. All parent-child heritability checks were as expected. Genotypes and B-allele frequency (BAF) data were exported using GenomeStudio v2.0.4 using the L1 version of the SNP manifest which is based on hg38.

For two individuals, for whom only whole genome sequencing was available, data was merged using PLINK v1.9^4^ and aligned to the positive strand using an established pipeline (https://www.well.ox.ac.uk/~wrayner/strand/index.html). The PLINK file contained 272,135 autosomal variants. Genetic maps were obtained from Beagle website (**https:// faculty.washington.edu/browning/beagle/beagle.html)** for hg38 and PLINK v1.9 was used to update the genotype map file with centiMorgan (cM) positions.

**Linkage analysis**

Variants with Mendelian errors identified by Pedstats v0.6.10^7^ were excluded. Mapthin v1.11 (<https://www.staff.ncl.ac.uk/richard.howey/mapthin/index.html>) trimmed the genetic data to ~5 SNPs/cM yielding 17,331 variants for linkage analysis. Merlin v1.1.2^8^ was used to perform the linkage analysis under a rare dominant model with a disease frequency of 0.001% and 100% penetrance. The extended chromosome 20 locus (chr20:46,200,000-53,700,000) was analysed using a harmonised set of build 38 variants that were present in all 3 families using the same rare dominant model and -*best* flag to haplotype and identify recombination events.

**UK Biobank haplotype frequency**
European ancestry UK Biobank participants (application 11223) best guess imputed genotypes were extracted for the linkage peak on chromosome 20 shared between families 1 & 2. The genotypes were loaded to Haploview v4.1 and all haplotypes were inspected for the risk haplotype above the frequency of 0.001%.

**Variant detection and annotation**

Using the six genomes sequenced, paired-end reads were aligned to the hg38 reference genome using bwa v0.7.15^9^ and duplicated reads were marked using samblaster v0.1.24^9^. Following quality checks, all samples resulted in median coverage of 26-39 X with ≥ 98 % of bases covered at least 10X (Supplemental Fig. 13 and Supplemental Table 6). Small variants were jointly called from individual BAM files using platypus v.0.7.9.5^10^ to generate a cohort VCF file representing variants detected in all six samples. In addition to standard platypus filters, we marked variants that do not have GQ ≥ 20 in at least one sample as “low quality”. The filtered VCF file was decomposed and normalized using vt v.0.57721^11^ and then annotated for protein-coding genes using SnpEFF v4.3^12^. Additional annotations were added using vcfanno v.0.3.1^13^. They included: i) allele frequencies from gnomAD v.3, 1000G phase 3, and an internal cohort of 300 WGS samples that were analysed using the same pipeline described above; ii) phyloP100 conservation score; iii) CADD v.1.5 score^14^. Finally, we annotated possible impacts on regulatory regions using GREEN-DB^15^, which includes a dataset of ~2.3 M regulatory regions with information on controlled genes and prediction scores specific to non-coding variants such as ReMM score v.0.3.1^16^.

**Selection of candidate variants**

Starting from the cohort dataset containing 6,926,029 variants we selected high confident heterozygous variants shared across the six genomes and never seen in the general population or our internal cohort. Confident heterozygous variants were selected with the following criteria: i) variant coverage ≥ 10 and variant GQ ≥ 20; ii) alternate allele-fraction between 0.2 and 0.8; iii) fraction of reads with low MQ < 0.20; iv) not located in a segmental duplication or low complexity region as previously defined^16^. The final pool of candidate variants was then selected as follows: i) absent in gnomAD v.3, 1000G phase, and the internal cohort; ii) heterozygous variant shared across all six individuals.

**Decoding of GWAS Signals**

GWAS variants were assessed using a multi-omics and machine learning platform as previously described^17^. 20q13.13 sentinel variants from a GWAS study for ST wave traits^18^ were tested for linkage disequilibrium (LD) using LDlink^19^ and two main signals identified (Supplemental Dataset 1). 81 variants in high LD (r^2^≥0.8, EUR) with sentinel variants were then assessed for potential coding, splicing and regulatory mechanisms. Putative coding changes were identified using ANNOVAR^20^ (v 20211019). Splicing changes for 17 intronic variants were assessed from the reported whole genome scan using SpliceAI (v1.3.1)^21^. Candidate regulatory variants were then identified using cardiomyocyte open chromatin data^22,23^ and deepHaem (model 5, https://github.com/rschwess/deepHaem/tree/master/ models)^24^ prediction of accessibility changes.

**Deep learning based variant effect prediction**

A convolutional neural network for predicting chromatin features^25^ was trained using a total of 936 publicly available chromatin feature data (DNase/ATAC-seq, transcription factor and histone modification ChIP-seq) as previously described^24^, and supplemented by cardiomyocyte and progenitor ATAC-seq data from GSE89895 (https://www.ncbi.nlm.nih.gov/ geo/query/acc.cgi?acc=GSE89895)^22^ and GSE106690 (https://www.ncbi.nlm.nih.gov/geo/ query/acc.cgi?acc=GSE106690)^23^. The impact of the indel variant on chromatin accessibility in cardiomyocytes was predicted as the differences between the cardiomyocyte accessibility probability model output from the 13 wild type 1 kb regions against the same region containing the variant sequence (chr20:49,356,363-49,357,375, hg38). For the *in silico* mutagenesis the effect of every possible single base pair substitute at each base pair was computed.

**Open chromatin assay**

ATAC-sequencing ^26^ was performed with 1.5 × 10^5^ cardiomyocyte cells which were lysed for 15 min in ice-cold lysis buffer [10 mM Tris-HCl pH 7.5, 10 mM NaCl, 3 mM MgCl_2_, 0.1% Igepal CA-630], before centrifugation at 500 rcf (15 min, 4ºC).The lysis buffer was then discarded and the DNA was transposed for 15 minutes at 37ºC (47.5 µL TD Buffer, 2.5 µL Tn5 Transposase [Illumina]). DNA was purified using MinElute (Qiagen) and amplified with custom indexed sequencing adaptors^26^. Libraries were sequenced on an Illumina NextSeq Platform using 40 bp paired-end reads and mapped to hg38 using NGSeqBasic^27^ (v20) with bowtie2 (v2.4.2). Unmapped reads were saved and then remapped to a custom genome containing the indel sequence.

**Cleavage Under Targets and Release Using Nuclease (CUT&RUN)**

CUT&RUN was carried out as described^28^. Briefly, Concanavalin A magnetic beads were activated by washing twice in binding buffer (20 mM HEPES-KOH pH 7.9, 10 mM KCl, 1 mM CaCl_2_, 1 mM MnCl_2_). 100,000 iPSC-derived cardiomyocytes were washed twice in 1.5 mL of wash buffer (20 mM HEPES pH 7.5, 150 mM NaCl, 0.5 mM spermidine and one Roche Complete Protease Inhibitor tablet) then resuspended in 200 μL wash buffer, mixed with 10 μL activated bead suspension and rotated for 5–10 min at room temp. Tubes were then placed in a magnetic rack, the wash buffer was replaced with 200 μL of antibody buffer (wash buffer plus 0.02% digitonin, 2 mM EDTA and 1 µL of either anti-H3K4me1 [1:200, AbCam; ab8895, lot GR3206285-1] or anti-H3K4me3 [1:200, Millipore; 07-473, lot 2664283]) and the tubes rotated for 2 hrs at 4°C. Tubes were then placed on a magnetic rack and the cells were washed once with digitonin buffer (wash buffer plus 0.02% digitonin) before being resuspended in 200 μL digitonin buffer. pA-MNase enzyme (a kind gift of the Henikoff lab) was added to each tube to a final concentration of 700 ng mL^-1^ and the tubes rotated for 1 hr at 4°C. Tubes were then placed on a magnetic rack and the cells were washed twice with digitonin buffer before being resuspended in 150 μL digitonin buffer. Digestion was initiated by the addition of 3 μL 100 mM CaCl_2_ to each sample and was allowed to proceed whilst tubes incubated on ice for 30 min before being quenched with 50 μL 4x stop buffer (680 mM NaCl, 40 mM EDTA, 8 mM EGTA, 0.04% digitonin, 0.1 mg mL^-1^ of RNase A, 0.1 mg mL^-1^ glycogen). Tubes were incubated for 10 min at 37°C, then centrifuged for 5 min at 4°C at 16,000 rcf. The supernatant was moved to fresh tubes, mixed with 1 μL of 20 % SDS and 1.5 μL of 20 mg mL^-1^ proteinase K, then incubated for 10 min at 70°C. DNA was purified by phenol-chloroform extraction and ethanol precipitation, then libraries were prepared using the Ultra II DNA library prep kit (NEB) following the manufacturer’s protocol with 13 PCR cycles. Libraries were sequenced with 150 bp paired-end reads by NovoGene and mapped to hg38 using NGSeqBasic^27^ (v20). To classify open chromatin regions as either putative promoters or putative enhancers ATAC-seq from cardiomyocytes containing the InDel were identified using the LanceOtron^29^ (v2) deep learning neural network. To capture surrounding chromatin signatures, peaks were extended in both direction by 1 kb using bedtools slop (v2.29.2). 68,236 extended peak calls and bigwigs for H3K4me1 and H3K4me3 CUT&RUN were uploaded into Multi Locus View^30^ where log(H3K4me1/H3K4me3) was calculated using density scores. Peaks with a negative score were assigned as putative promoters (n=23,361; 34.2 %), peaks scoring 0 remained undetermined (n=2,538; 3.7 %), and peaks scoring >0 were annotated as putative enhancers (n=42,337; 62.0 %).

**Chromosome conformation capture**

Processed Hi-C data for RUES2 and WTC-11 derived cardiomyocytes^23^ (GSE106687) were downloaded, valid Hi-C interaction pairs were converted to contact matrices and normalised using iterative correction and eigenvector (ICE) decomposition with HiC-Pro^31,32^ (v2.7.5b). NuTi capture-C experiment was performed as previously described^33^. Briefly, 1-5 x 10^6^ cells were first lysed on ice in 5 mL lysis buffer and then pelleted by centrifugation (15 min, 4ºC, 500 rcf). The lysis buffer was then discarded and the nuclei were resuspended in 1 mL PBS before snap freezing with dry ice and ethanol. Later, the nuclei were defrosted, pelleted (15 min, 4 ºC, 500 rcf) and resuspended in 215 µL 1× *Dpn*II buffer. They were then permeabilized with 0.28 % SDS for 1 hour at 37ºC (200 µL nuclei, 60 µL 10× *Dpn*II buffer, 434 mL PCR grade water, 10 µL 20 % vol/vol SDS) and the SDS was quenched by addition of 66 µL of 1.67 % Triton-X. Next, *Dpn*II was added in three aliquots of 10 µl (500 U) several hours apart for a total digest time of 20 hours at 37ºC. The enzyme mixture was then heat inactivated at 65 ºC for 15 min and immediately cooled on ice. The digested DNA was then ligated by addition of 240 U T7 ligase (500 mL PCR grade water, 134 mL 10× ligation buffer, 8 µL ligase) and incubated overnight at 16ºC. Following ligation, nuclei were isolated by centrifugation (15 min, 4 ºC, 500 rcf), resuspended in 300 µL of TRIS-EDTA, and de-crosslinked overnight at 65 ºC with 5 µL Proteinase-K (3 U). RNA was then removed from the mixture by addition of 5 µL RNAse A (7.5 mU) for 30 min at 37 ºC. Next, the DNA was extracted by addition of 310 µL phenol-chloroform-isoamyl alcohol, centrifugation in a phase lock tube (10 min, 12,600 rcf, room temp), and overnight ethanol precipitation at -20 ºC (30 µL 3 M sodium acetate, 1 µL glycoblue, 900 µL 100 % ethanol). Following ethanol precipitation, the DNA was pelleted by centrifugation (30 min, 21,000 rcf, 4ºC) and washed twice with 70 % ice cold ethanol before resuspension in 150 µL water (30 µL for controls).

3C libraries were sonicated on a Covaris S220 to 200 bp and 2 µg of sonicated DNA was indexed with NEBNext Ultra II DNA Library Prep Kit for Illumina (New England Biolabs) with the following modifications: for the End Prep reaction, the 20 ºC incubation was lengthened to 45 min, 5 µL of NEBNext Adaptor was added and incubated for 30 minutes at 20 ºC, the USER Enzyme incubation was extended to 30 minutes (37 ºC), and indexing was performed in two reactions with Herculase II Fusion Polymerase (Agilent) using six cycles of amplification. For the capture experiments, two separate pools of 70-mer biotinylated oligonucleotides (Sigma) were combined to target either promoters or enhancers (Supplemental Table 7). Oligonucleotide pull-down assays for single and double capture of multiplexed 3C libraries were performed using the Nimblegen SeqCap EZ kit (Roche) following manufacturer’s instructions using a single reaction per library for the primary capture and a single capture per pool for the double capture. Oligonucleotides were used at a concentration of 23.2 nM and the DNA was amplified with 10 PCR cycles. Prepared libraries were then sequenced on an Illumina MiSeq platform with 150 bp paired-end reads. Reads generated were then mapped to hg38 using CCseqBasic^34^ (v5) with bowtie2 (v2.4.2) using default settings. Final interaction profiles were then windowed using CaptureCompare^34^ (v1) with 200 bp bins and 4 kb windows.

**Zebrafish enhancer cloning**

Enhancer sequences (Supplemental Table 8) were generated as custom-made, double-stranded linear DNA fragments (GeneArt® Strings™, Life Technologies). The DNA fragments were then cloned into the pCR8 vector using the pCR8/GW/TOPO TA Cloning Kit (Invitrogen, K2500-20) following manufacturer’s instructions. The enhancer inserts, from positively cloned constructs, were transferred from the pCR8/GW/enhancer entry vector to a suitable destination vector using Gateway LR Clonase II Enzyme mix (Life Technologies, 11791-100) following manufacturer’s instructions. For transgenesis, the enhancers were cloned into the E1b-GFP-Tol2 vector.

**Zebrafish maintenance and image analysis**

All zebrafish procedures were performed in compliance with ethical regulation, approved by the Clinical Medicine Local Ethical Review Committee at University of Oxford, and licensed by the UK Home Office (PP1224162). Mosaic transgenic zebrafish embryos were generated from *tg(myl7:H2A-mCherry)* zebrafish^35^ using Tol2 mediated integration^36^. Embryos were maintained in E3 medium (5 mM NaCl; 0.17 mM KCl; 0.33 mM CaCl2; 0.33 mM MgSO4) at 28.5 °C. For image analysis, F0 transgenic zebrafish embryos were anesthetized with 0.05 % tricaine methanesulfonate (Sigma-Aldrich) and single embryos were transferred into flat bottom 96-well plates that were mounted in topvision low melting point agarose (ThermoFisher Scientific). GFP and mCherry reporter gene expression was screened with a Zeiss LSM 710 confocal microscope at 5 dpf. Hearts of zebrafish embryos were imaged using the ZEN Black 2012 SP1 software using the tile scan command, combined with Z-stack collection under a Zeiss LSM 710 MP (Carl Zeiss) confocal microscope at 488 nm excitation and 509 nm emission (EGFP) and 587 nm excitation and 610 nm emission (mCherry). Image analysis was performed using Fiji (ImageJ 1.53f).

**Human iPSC line gene-editing with CRISPR/Cas9**

HPSI0114i-Kolf_2 human iPS cells (HipSci, cat# 77650100) were cultured at 37 °C with 5 % (v/v) CO_2_ on vitronectin coated plates (Thermo Fisher cat# A31804) in Essential 8 medium (Thermo Fisher cat# A1517001). For single cell preparations, cells were dissociated using TrypLE (Thermo Fisher cat# 12563011) and maintained in E8 medium containing either 10 uM ROCK inhibitor or 1X RevitaCell supplement (Thermo Fisher cat# A2644501).

A sgRNA targeting the sequence of 5’-CCCTCCTTGGGGAGAGGAAC-3’, close to the delinsTCCC site, was designed using the CRISPOR algorithm (http://crispor.tefor.net/) and synthesized (Synthego). A 139-nucleotide homology repair template (5’-TTGCTGTGGGG CCATGTGATCATCAGGAGGGATAGGTCCTCAAACAGGCGATGAGCTCAGGACATGGGCATTTCCCCCCCAAGGAGGGGGGAAAGCTGTGCAGTCACAGACAACGTGGCCTTTTATAAGCTTGAATGTGATGT-3’) (Eurogentec) was designed to introduce the 17 base pair deletion and insertion of TCCC which ablates a *Bsr*DI restriction enzyme site. A ribonucleoprotein complex was prepared using 1 µl synthetic sgRNA (20 µM) and 0.6 µl Engen Spy Cas9-NLS (20 µM, NEB cat# M0646T) in 4.4 µl Neon transfection buffer R following the manufacturer’s protocol. 10^5^ cells were electroporated with the RNP complex and ssODN (300 nM) using the Neon transfection system (1200 volts, 30 ms, 2 pulses). Individual clones were genotyped using a PCR designed to recognize the mutant, but not the wild-type allele, amplifying a 293 bp sequence from the correctly targeted allele (5’-TTGTGATTCACGTTCCCTCTCC-3’, 5’-TTCCCCCCTCCTTGGGGGGGA-3’). The loss of a *Bsr*DI restriction site, in correctly targeted alleles, was confirmed by digesting PCR amplicons (generated using primers 5’-TTGTGATTCACGTTCCCTCTCC-3’, 5’- CTCTGATCCCTGTCAGCCTTG-3’) spanning the targeted region with *Bsr*DI (NEB cat# R0574S). Editing was confirmed through Sanger sequencing of this latter amplicon. All the clones were quality controlled by assaying for pluripotent stem cell transcription factor expression using FACS (BD cat# 560589) and a normal karyotype was confirmed by staining of metaphase chromosome spreads using standard techniques.

**Human iPSC culture of gene-edited and wild type control lines**

Following gene-editing, continued culturing of iPSC lines took place at 37 °C with 5 % (v/v) CO_2_ in E8 medium (TeSR™-E8™, Stemcell Technologies) on feeder-free plates coated with matrigel hESC-qualified matrix (Corning). Cells were passaged at a 1:4 ratio by dissociation with TrypLE^TM^Select Enzyme (1X) (Gibco) followed by culturing for 24 hours in E8 medium supplemented with 10 µM Rho kinase inhibitor Y-27632 (Abcam). After 24 hours, the medium was replaced with standard E8 medium and changed daily till cells reached 80 % confluency (2-4 days).

**Cardiomyocyte culture and differentiation**

The iPSC lines were differentiated into cardiomyocytes using protocols adapted from Lian et al.^37^ and Zhang et al.^38^. Briefly, cells were seeded at a 1:5 ratio (approximately 200,000 cells/well) on 24 well plates coated 1:50 with reduced factor matrigel (matrigel basement membrane matrix growth factor reduced LDEV-free, Corning). They were then cultured in E8 medium until reaching 90 % confluency (day 0).  On day 0, -INS medium (RPMI 1640, Life Technologies Ltd, with N21-MAX insulin free media supplement, Bio-Techne) containing 8 µM CHIR-99021 (Stratech Scientific Ltd), 10ng/ml recombinant Activin A (Bio-Techne), and 1:100 reduced factor matrigel were added for 24 hours. Following incubation (day 1), the medium was replaced with -INS medium and cells were cultured for a further 48 hours until day 3. They were then cultured in 1:1 old to fresh -INS medium supplemented with 5 uM WNT inhibitor IWP-4 (Stemcell Technologies) until day 5, when the medium was replaced again with -INS medium for 48 hours. On day 7, the medium was switched to +INS medium (RPMI 1640, Life Technologies Ltd, with N21-MAX media supplement, Bio-Techne) and replaced every second day until more than 60 % of cells were beating (day 12-15). They then received daily medium changes until harvesting (day 21 for ATAC sequencing and day 38 for RNA and Capture-C experiments).

**Determining phased haplotypes for CRISPR/Cas9 edited iPSC**

BAM files generated from nanopore sequencing of the CRISPR/Cas9 edited iPSC were filtered to select reads matching phased data from the iPSC (HipSci, cat# 77650100). This was done by selecting reads with CTAT or GGGC haplotypes for SNPs rs2426151, rs3091764, rs6095492 and rs3092378. The CTAT haplotype for these cells is in cis with the coding SNP rs2229006-C allele, while GGGC is in cis with the rs2229006-G allele. Filtering was done using a custom Python script (Python 3.86), and the resulting files were then indexed using Samtools 1.4.1. The Integrative Genomics Viewer (IGV)^39^ version 2.8.10, was used to determine which haplotype and corresponding coding allele from rs2229006 was in cis with the delinsTCCC variant.

**RNA extractions and cDNA synthesis**

Messenger RNA was extracted from cardiomyocytes using the Direct-zol RNA Miniprep kit (Zymo Research catalogue no. R2051) according to manufacturer’s instructions. For quantitative PCR experiments, 1 µg RNA was converted into cDNA using the SuperScript® III First-Strand Synthesis System (Invitrogen).

**Quantitative PCR**

Real-time quantitative PCR amplification were performed in duplicate for four differentiations of the wild type (C1) and heterozygous clones (C6, C68 and C93). A single sample, for the C6 clone, failed to amplify. All reactions were set up using 8 µl cDNA, 10 µl TaqMan™ Fast Universal PCR Master Mix (cat no. 4352042, Applied Biosystems), 1 µl KCNB1 FAM-MGB TaqMan assay (Hs00270657_m1) and 1 µl GAPDH VIC-MGB TaqMan assay (Hs02758991_g1). PCR amplifications were performed with a fast QuantStudio 7 Flex system cycling at 95 °C for 20 seconds followed by 65 cycles of 95 °C for one second leading into 60 °C for 20 seconds. Cycle threshold (CT) expression values were recorded and normalised to GAPDH as an internal, housekeeping control in a comparative CT analysis. As the data for the heterozygous clones was not normally distributed (Shapiro-Wilk test, p= 0.0148), the non-parametric Mann-Whitney statistical test was used for analysis.

**RNA sequencing**

Messenger RNA extracted from day 38 cardiomyocyte differentiations (n=3) of one of the heterozygous clones (C6) was investigated further by RNA sequencing. Sequencing was completed by the Oxford Genomics Centre at the Wellcome Trust Centre of Human Genetics, selecting Poly+ mRNA and generating double stranded libraries using NEBNext Poly(A) mRNA Magnetic Isolation Module (E7490) and NEBNext Ultra II Directional RNA Library Prep Kits form Illumina (E7760L) with specific Illumina adapters and barcode tags (dual indexing^40^). The RNA-sequencing libraries were pooled and sequenced on an Illumina NovaSeq6000 instrument as 150 bp paired end reads (approximately 1.85x10^8^ per sample). The data collected was aligned to the human reference genome (GRh37/hg19) and quality checked, fastq files were processed with Phred+33. Allele-specific expression analysis was performed using Integrative Genomics Viewer (IGV)^39^ version 2.8.10.

**Data availability**

Capture-C (Figure 7 and Supplementary Figure 11), ATAC-seq (Figures 4,5,7, Supplementary Figures 7, 11), CUT&RUN (Figure 5, Supplemental Figure 8) and RNA-seq (Supplementary table 5) raw and processed sequencing data are available on the Gene Expression Omnibus (GSE163174).

To review GEO accession GSE163174:

Go to https://www.ncbi.nlm.nih.gov/geo/query/acc.cgi?acc=GSE163174

Enter token yvkpomiarrqlpol into the box

**Code availability**

Custom analysis codes are available on GitHub (https://github.com/BSGOxford/RTA-ST-syndrome-KCNB1), as are deepHaem model 5 (https://github.com/rschwess/deepHaem), CCseqBasic5 (github.com/Hughes-Genome-Group/CCseqBasic5) and captureCompare (github.com/Hughes-Genome-Group/CaptureCompare).

**Public Datasets analyzed**

Publicly available ATAC-seq, ChIP-seq data and Hi-C data (Figures 4,5,7 Supplementary Figures 4) was sourced from the Gene Expression Omnibus (GSE89895, GSE106690, GSE125926) and the Descartes Atlas (https://descartes.brotmanbaty.org/bbi/human-chromatin-during-development/).
