## Supplementary material for "An autosomal dominant cardiac arrhythmia syndrome, ST Depression Syndrome, is caused by the *de novo* creation of a cardiomyocyte enhancer": Supp Figures and Tables

**Supplementary Figures**

Supp. Fig. 1.	Haplotype analysis using Merlin software in Families 1 & 2	pg. 1-2

Supp. Fig. 2.	Frequency of risk haplotype in UK Biobank	pg. 3

Supp. Fig. 3.	*KCNB1* locus accessibility in developing heart and brain	pg. 4

Supp. Fig. 4.	Cell type specific activity of the wild-type sequence altered by delinsTCCC.	pg. 5

Supp. Fig. 5.	Machine learning prediction of the *de novo* activity of delinsTCCC sequence.	pg. 6

Supp. Fig. 6.	Quality control tests for heterozygous iPSC lines generated

with CRISPR-Cas9 editing	pg. 7

Supp. Fig. 7.	delinsTCCC ATAC-seq reads from heterozygous cardiomyocytes	pg. 8

Supp. Fig. 8.	Histone H3 modifications of cardiomyocyte open chromatin	pg. 9

Supp. Fig. 9.	Autofluorescence in zebrafish embryos	pg.10

Supp. Fig. 10.	Enhancer element E-139 expression in the brain of zebrafish embryos	pg.11

Supp. Fig. 11.	NuTi Capture-C from the *ZNFX1* and *PTGIS* promoter in cardiomyocytes	pg.12

Supp. Fig. 12.	Nanopore sequencing and phasing for the delinsTCCC variant

and the rs2229006 SNP	pg.13

Supp. Fig.13.	Sequence coverage for the individual genomes investigated	pg.14

**Supplementary Tables**

Supp. Table 1.	Linkage analysis results for Family 1 and Family 2.	pg.15

Supp. Table 2.	Variants shared across the six affected individuals	pg.16

Supp. Table 3.	Linkage analyses combined showing the interval with the delinsTCCC	pg.17

Supp. Table 4.	GFP expression in the heart of transgenic zebrafish	pg.18

Supp. Table 5.	Allele specific expression using RNA sequencing	pg.18

Supp. Table 6.	Summary quality metrics for WGS	pg.18

Supp. Table 7.	Chromosome conformation capture oligonucleotides	pg.19

Supp. Table 8.	Enhancer sequences for transgenic zebrafish	pg.20-21

**SUPPLEMENTARY FIGURES**

**Supplementary Figure 1.**

a)

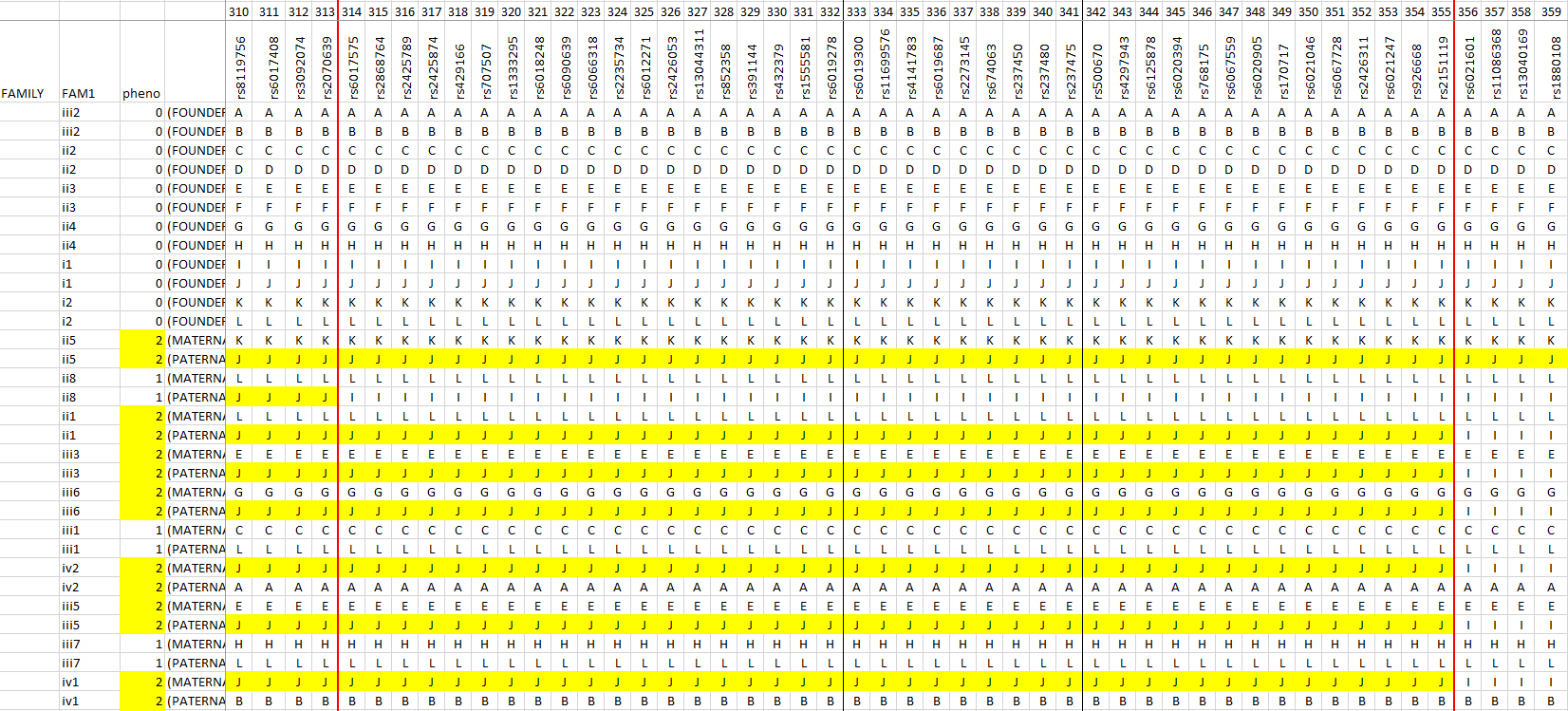

b)

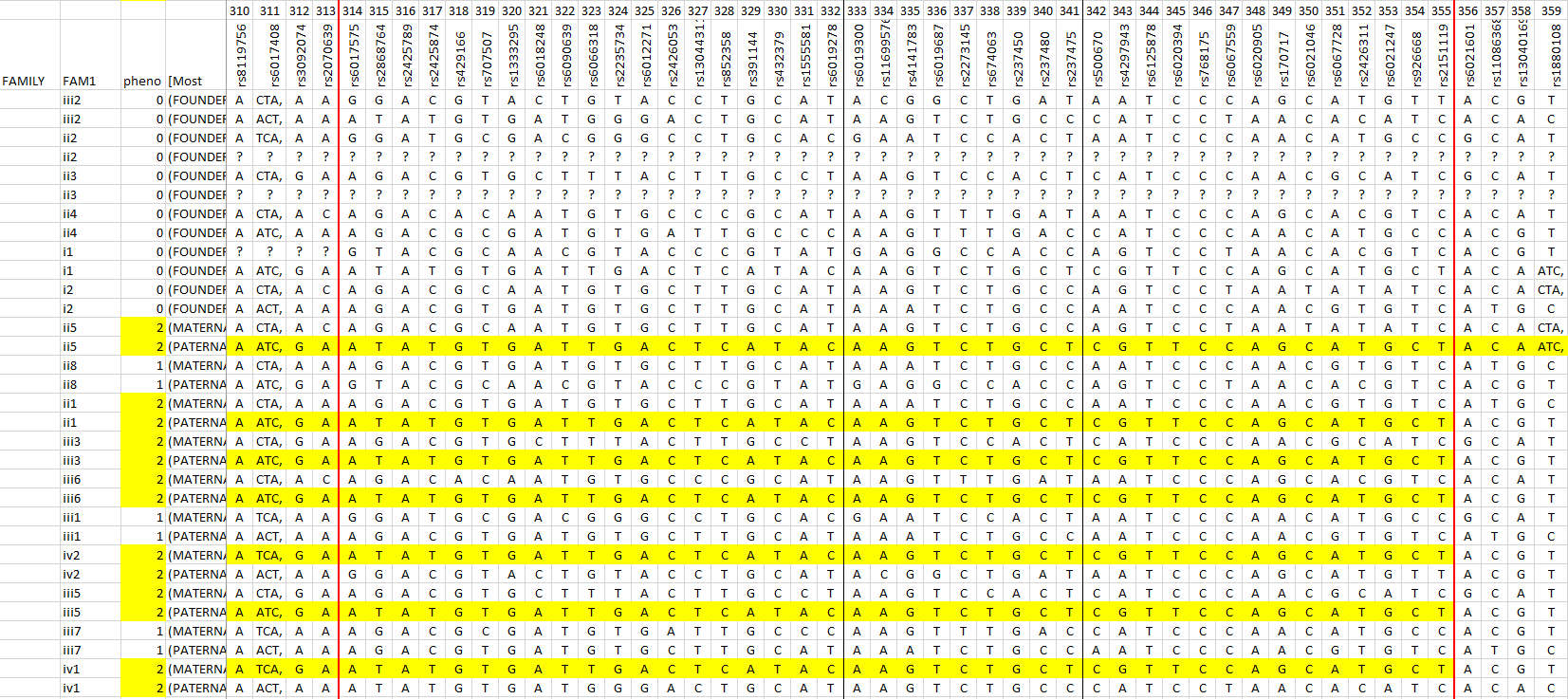

c)

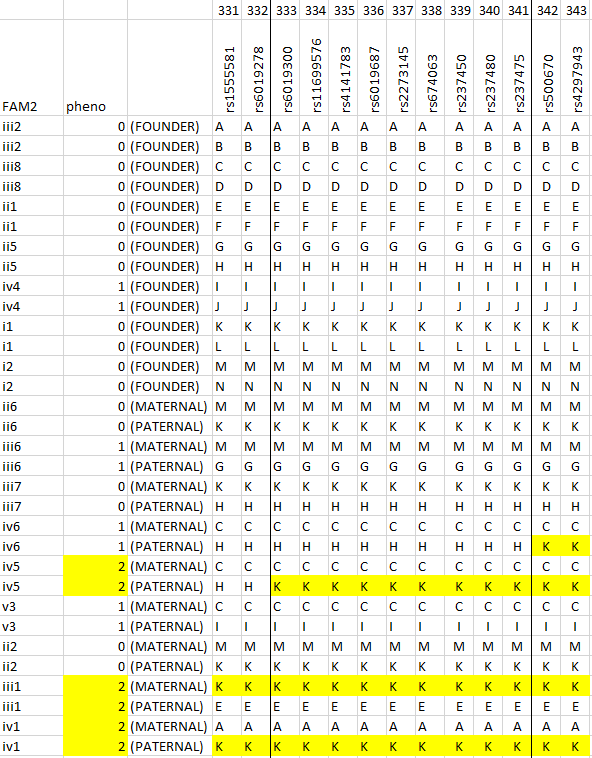

d)

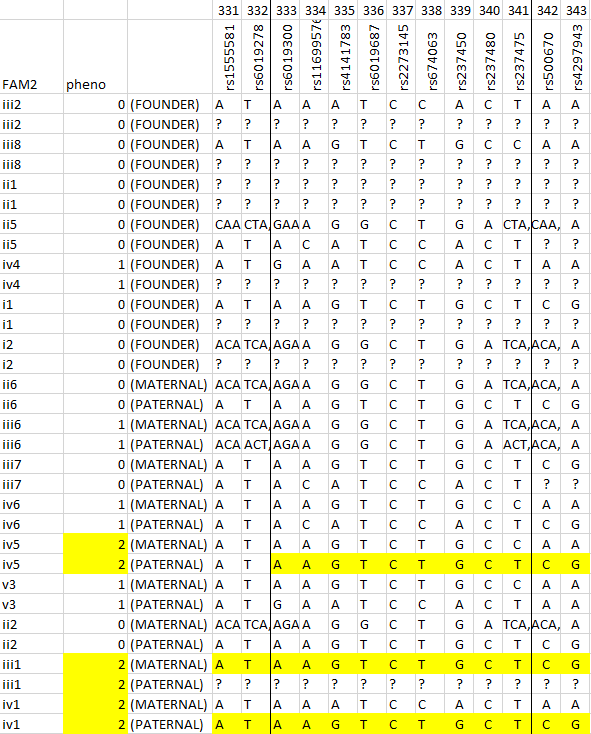

**Supp Fig 1: Haplotype analysis using Merlin software in Families 1 & 2 showing the most likely pattern of gene flow.** Panels a) & b) show the risk haplotype in yellow and the boundaries based on recombination events in Family 1 (red) and in Family 2 (black) marked by vertical lines; Panels c) & d) show the same for Family 2. The linkage region in Family 2 between markers 333-341 is indicated by black vertical lines and is also highlighted in a) & b) for ease of comparison.

**Supplementary Figure 2.**

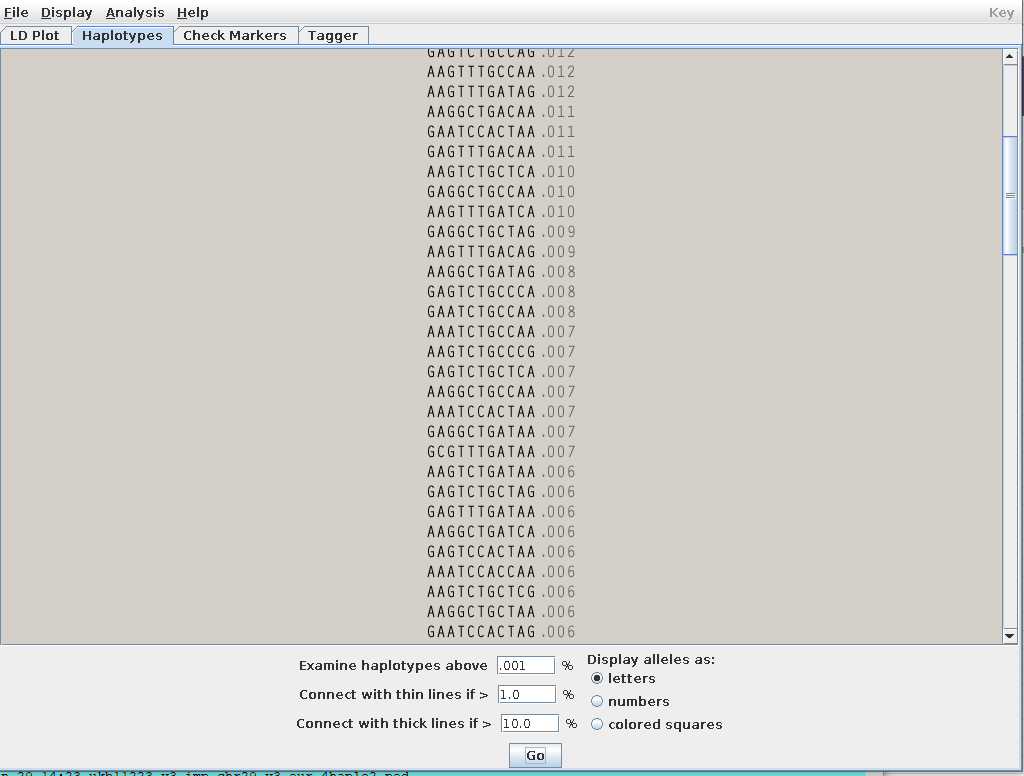

**Supp Fig 2: Frequency of risk haplotype in UK Biobank (based on 400K European participants)**

**Supplementary Figure 3.**

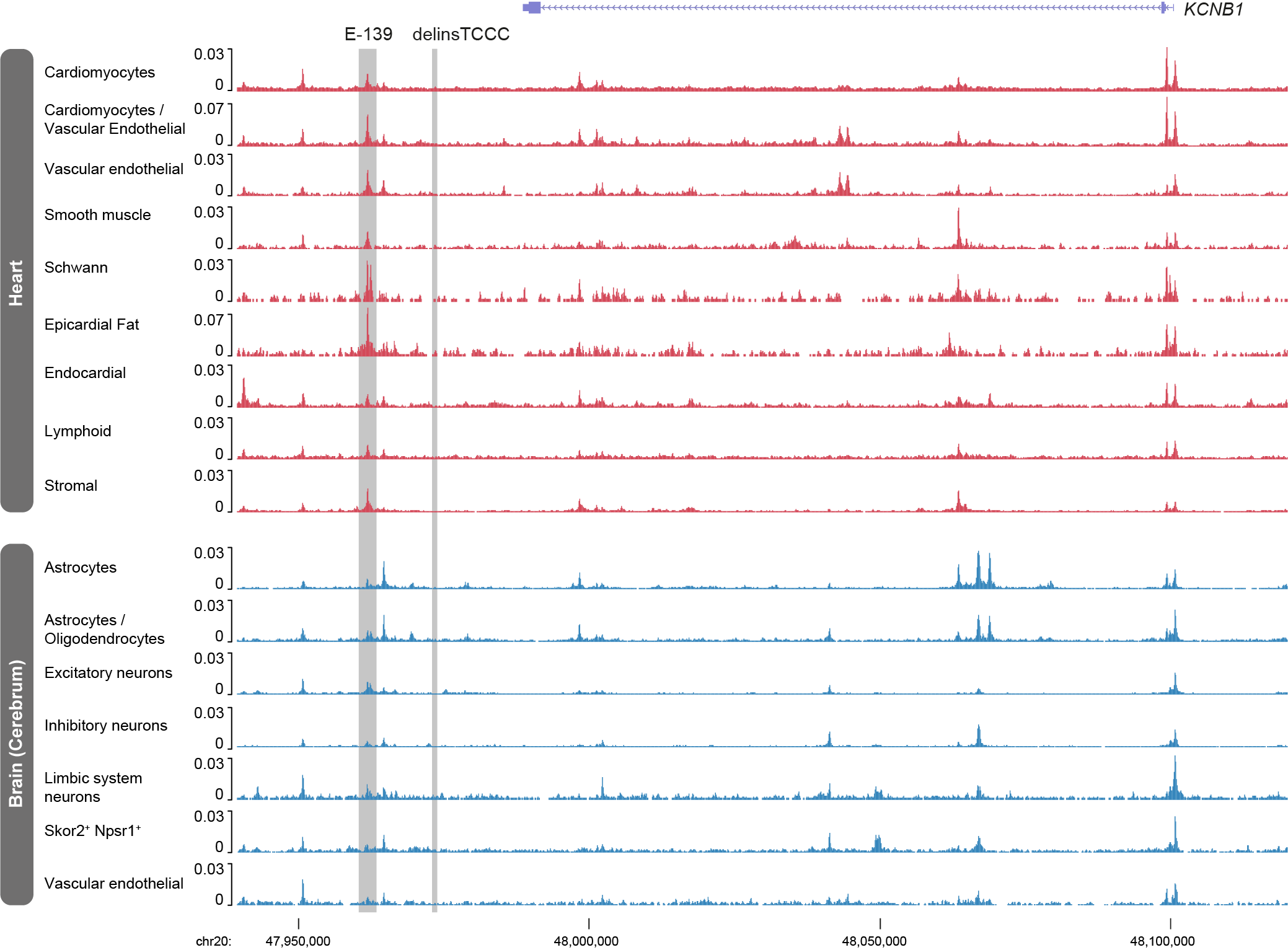

**Supp. Fig. 3. *KCNB1* locus accessibility in developing heart and brain.** Single-cell ATAC-seq from embryonic heart and brain tissues (Domcke *et al*. 2020). E-139 is a putative *KCNB1* enhancer (139 kb from the promoter and nearby to the delinsTCCC variant) that is present in both cardiac and brain cells. Region shown: chr20:47,940,000-48,120,000 (hg19).

**Supplementary Figure 4.**

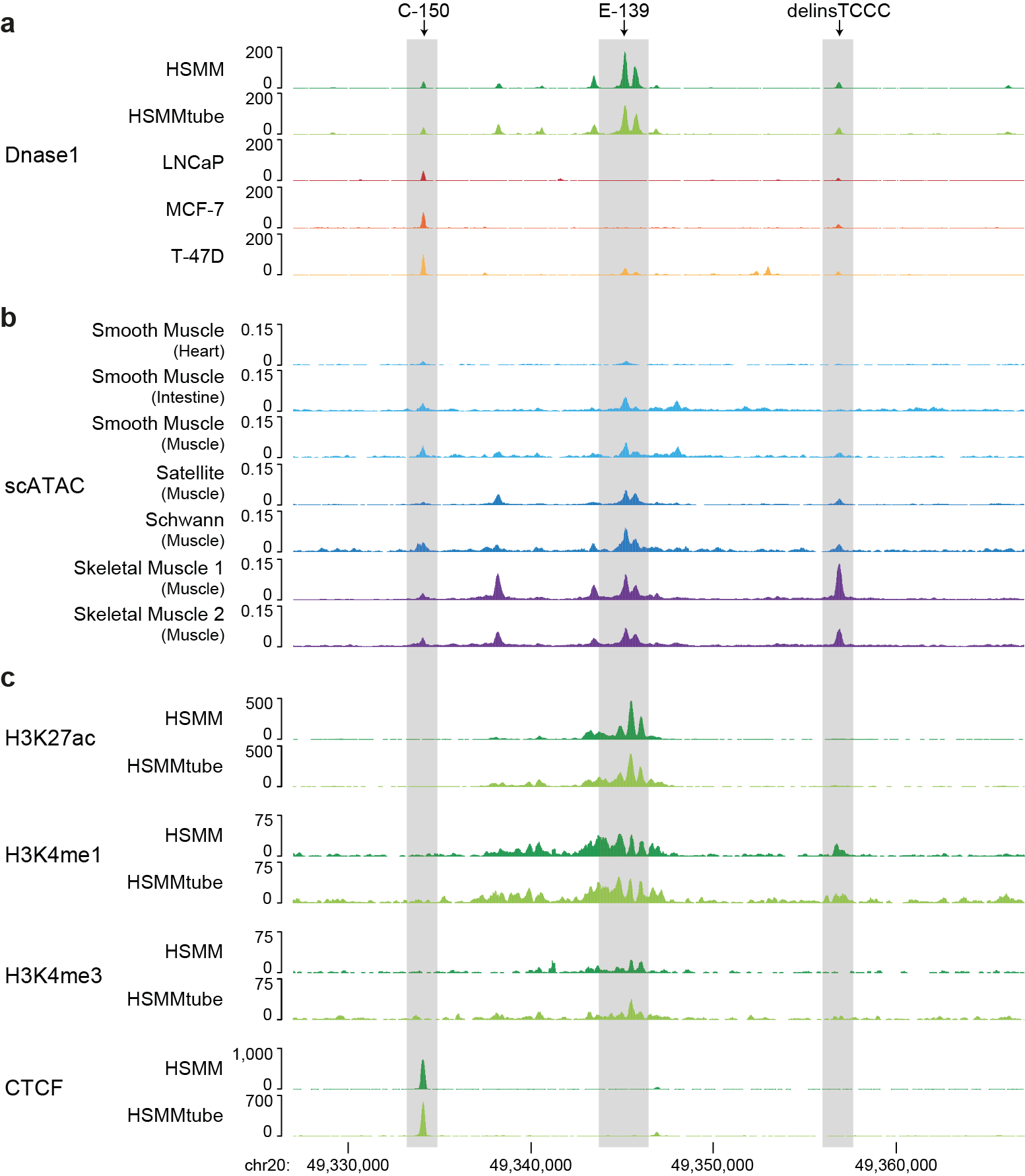

**Supp. Fig. 4. Cell type specific activity of the wild-type sequence altered by delinsTCCC.** In all panels the position of the CTCF bound region (C-150), the muscle enhancer (E-139) and the position of the wild-type sequence of delinsTCCC are named and high-lighted by grey bars in the various datasets. Panel A, shows cells with detectable open chromatin activity (DNase-seq, ENCODE) of the wild-type delinsTCC sequence in normal human skeletal muscle myoblasts (HSMM), normal human skeletal muscle myotubes (HSMMtube), prostate adenocarcinoma cell line (LNCaP), breast cancer adenocarcinoma cell line (MCF-7) and breast epithelial carcinoma cell line (T-47D). Panel B, shows cell types with detectable open chromatin activity (scATAC-seq) from a survey of 126 primary cell types during development. Panel C. the chromatin profile of the wild-type region in HSMM and HSMMtube cells for the enhancer associated chromatin marks (H3K4me1 and H3K27ac), promoter associated marks (H3K4me3 and H3K27ac) and for CTCF binding.

**Supplementary Figure 5.**

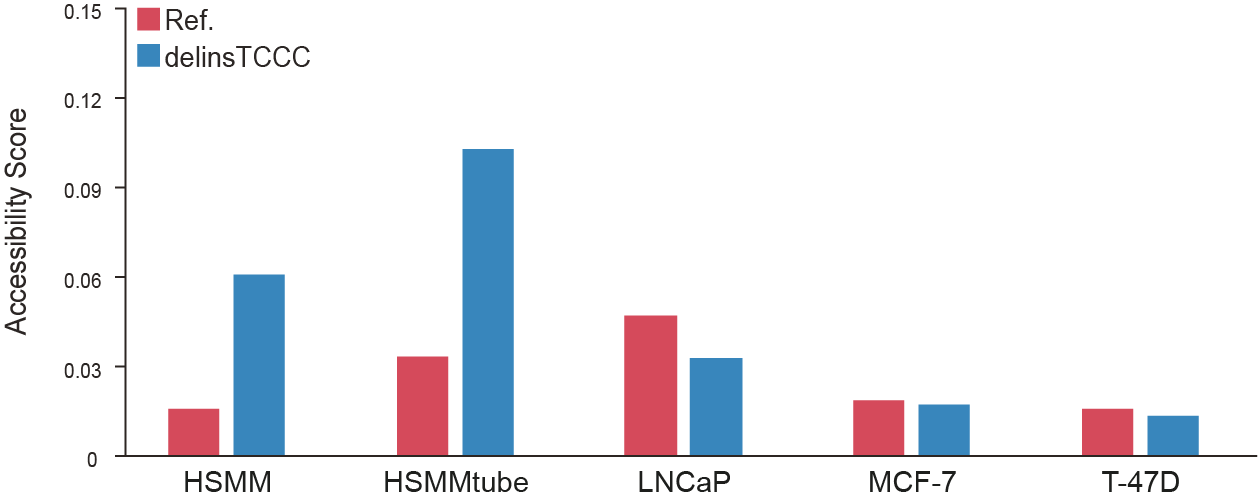

**Supp. Fig. 5. Machine learning prediction of the *de novo* activity of delinsTCCC sequence.** The Y axis show the relative predicted Accessibility score from the Convolutional Neural Network (CNN, see methods), trained on ENCODE chromatin datasets. The score for each cell type in Supp Fig 3 Panel A is shown for the reference sequence (red) and the sequence with the delinsTCCC mutation (blue).

**Supplementary Figure 6.**

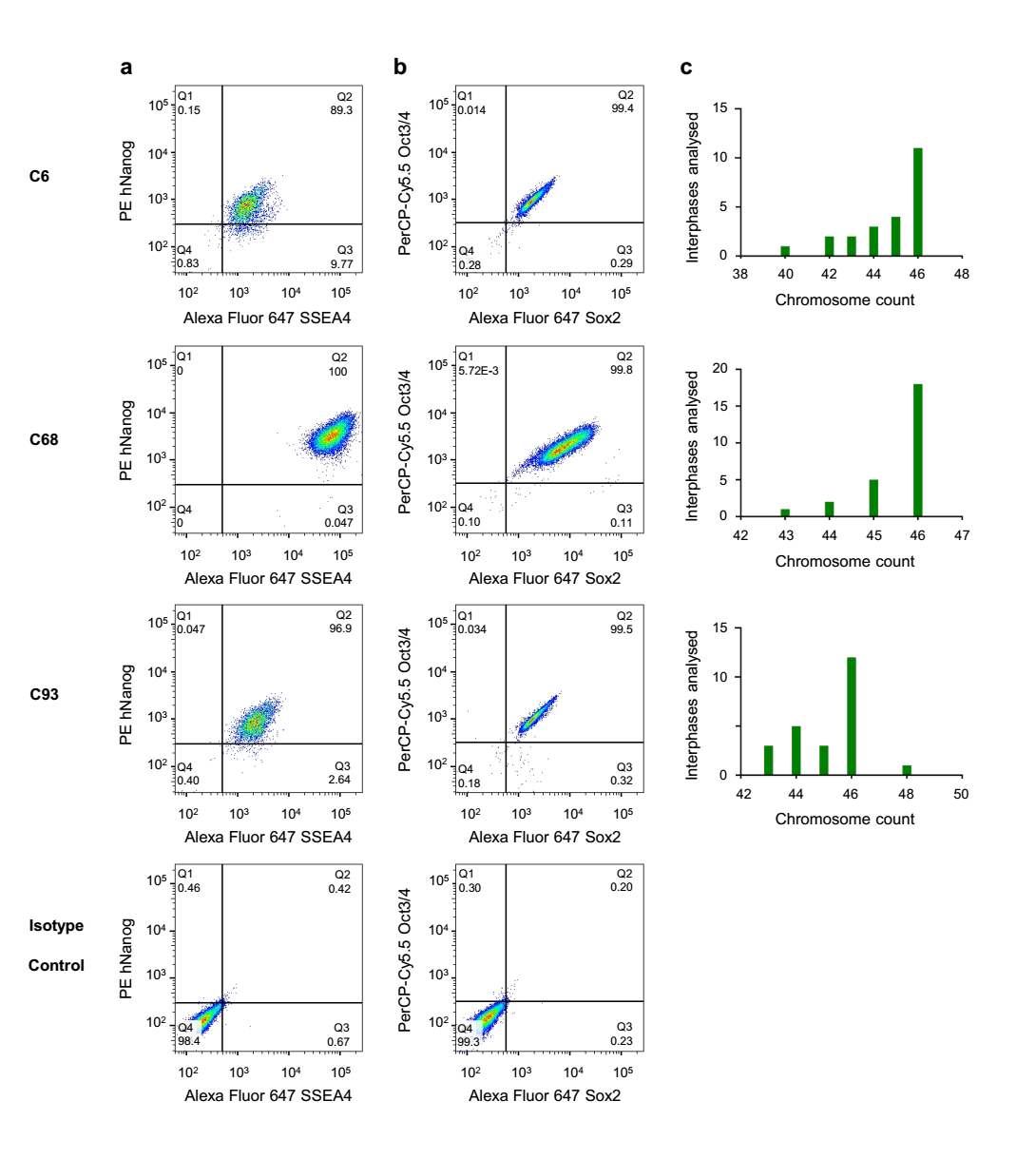

**Supp. Fig. 6. Quality control tests for heterozygous iPSC lines generated with CRISPR-Cas9 editing. a,** Flow cytometry analysis of pluripotent stem cell markers NANOG and SSEA4. The FACS analysis shows that more than 90% of cells are positive for both markers in clones C6, C68 and C93. **b**, FACS analysis for OCT4 and SOX2 shows more than 95% of cells are positive for both markers in all three clones. **c,** Karyotyping through analysis of metaphase spreads showed the modal number of chromosomes to be 46, indicative of predominantly diploid cells following CRISPR-editing and monoclonal expansion.

**Supplementary Figure 7.**

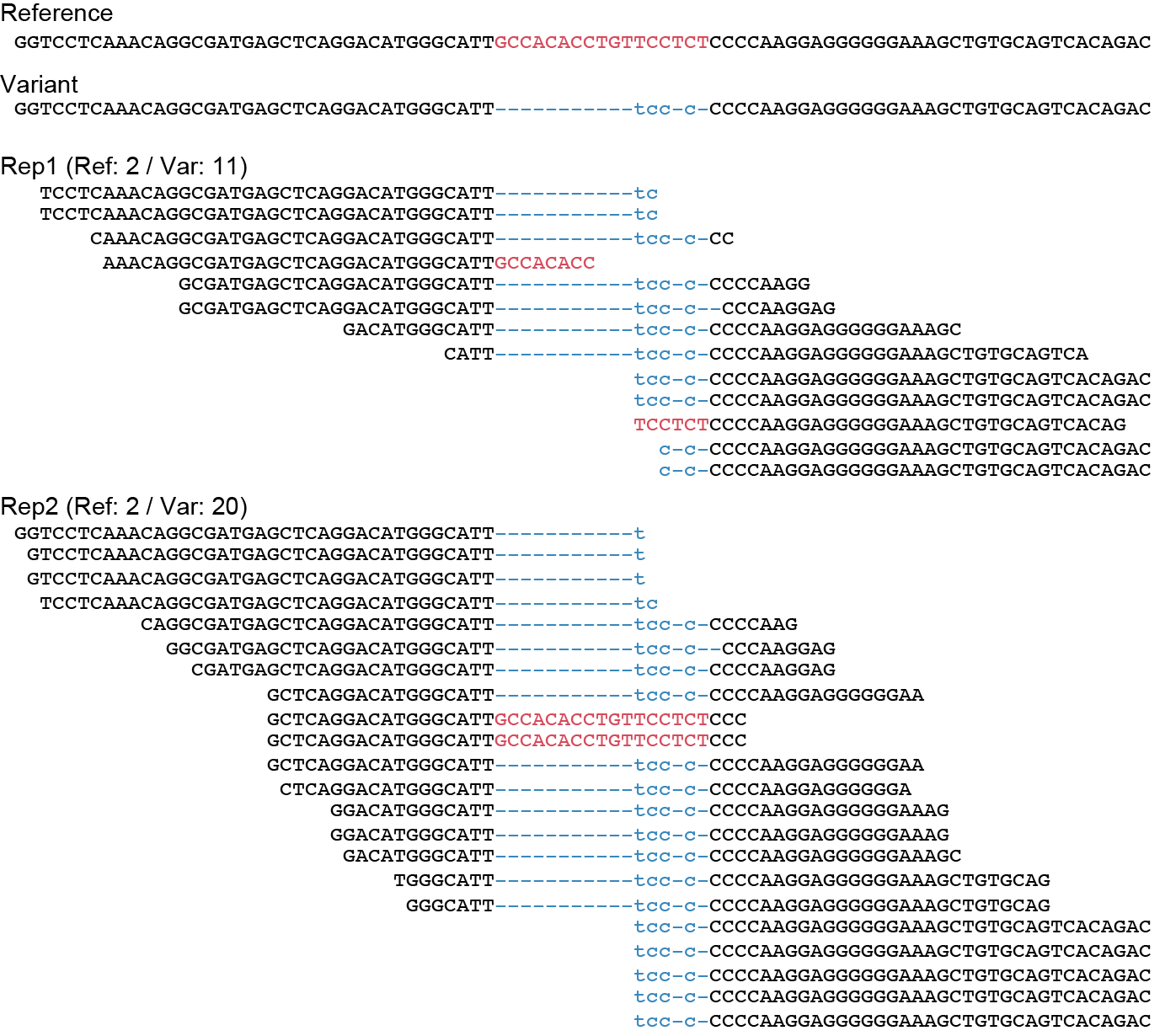

**Supp. Fig. 7. delinsTCCC ATAC-seq reads from heterozygous cardiomyocytes.** Reference (Ref) and delinsTCCC variant (Var) sequences with sorted unique ATAC-seq reads from cardiomyocytes generated from heterozygous iPSC cells. Paired-end reads were mapped first to the reference genome, unmapped read pairs were then mapped to a custom genome containing the delinsTCCC sequence, and PCR duplicates filtered.

**Supplementary Figure 8.**

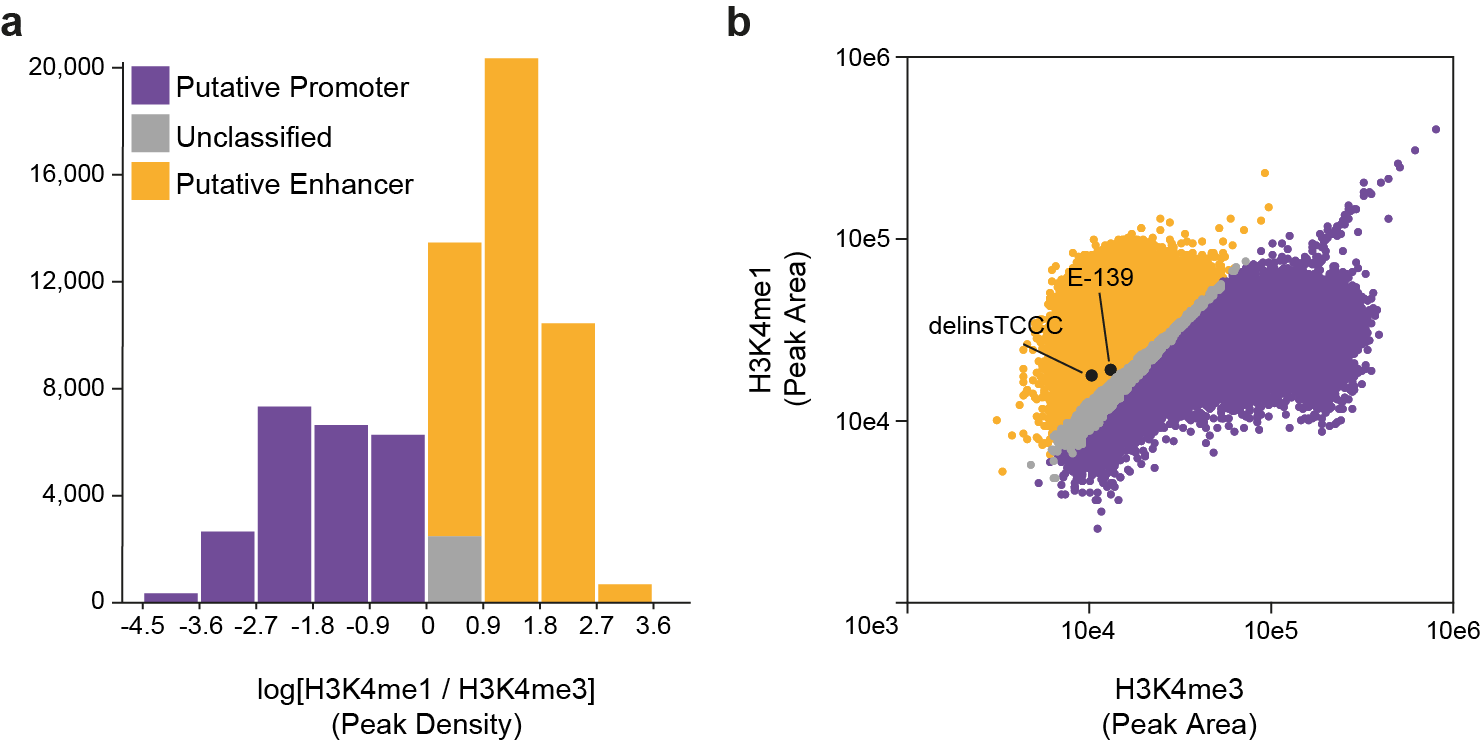

**Supp. Fig. 8. Histone H3 modifications of cardiomyocyte open chromatin. a,** Histogram showing distribution of CUT&RUN for H3K4me1 to H3K4me3 ratio values in cardiomyocytes generated from iPSC for regions of open chromatin identified by ATAC-seq (±1 kb). ATAC-seq peaks with a negative score were assigned as putative promoters (n=23,361; 34.2%), peaks scoring 0 remained unclassified (n=2,538; 3.7%), and peaks scoring >0 were annotated as putative enhancers (n=42,337; 62.0%). **b,** Scatterplot of ATAC-seq peak area scores for H3K4me1 and H3K4me3 CUT&RUN in cardiomyocytes with the delinsTCCC containing enhancer and the nearby putative enhancer (E-139) shown.

**Supplementary Figure 9.**

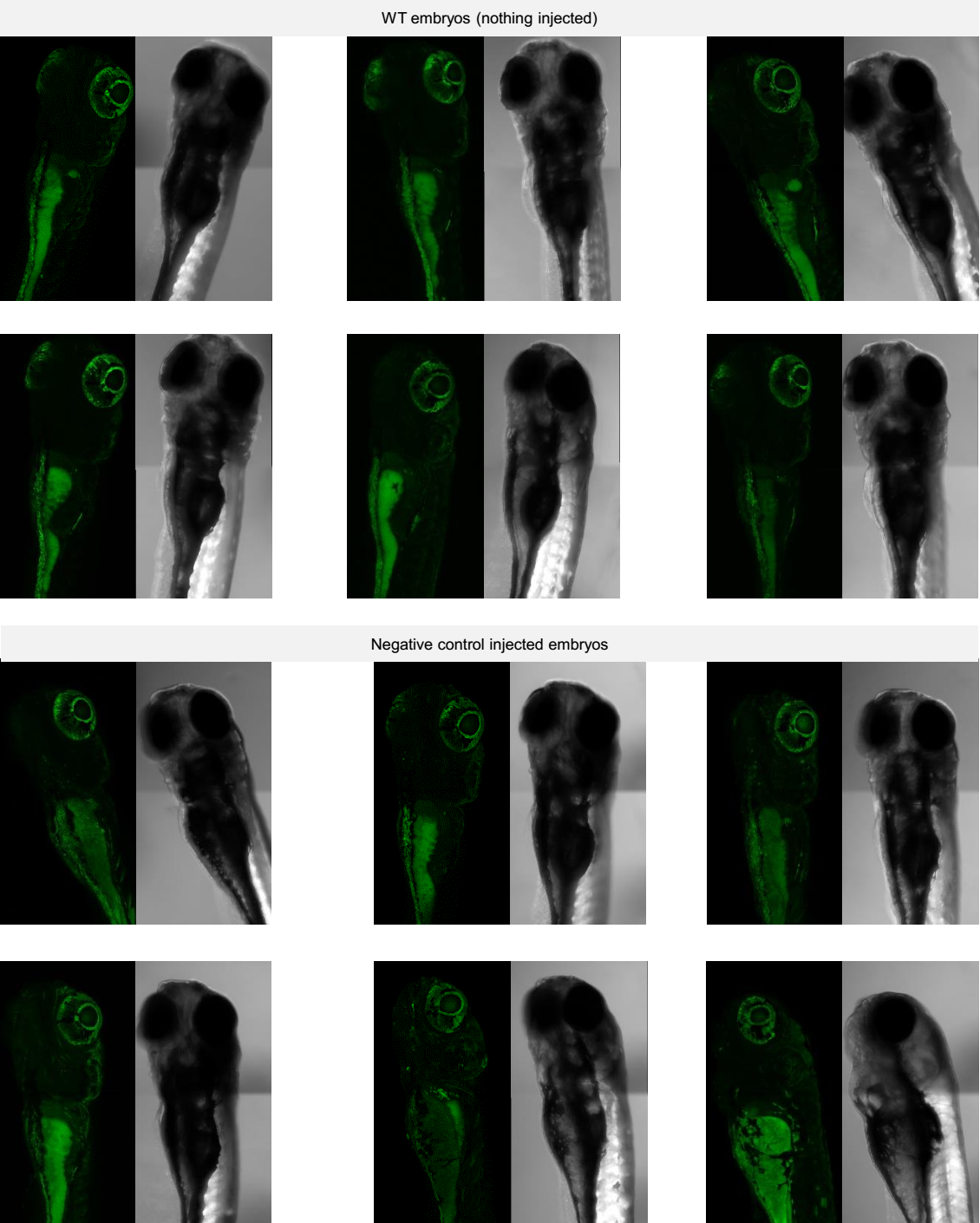

**Supp. Fig. 9. Autofluorescence in zebrafish embryos.** Zebrafish embryos at 5dpf without injections or negative control injections containing a random DNA sequence for delinsTCCC variant site.

**Supplementary Figure 10.**

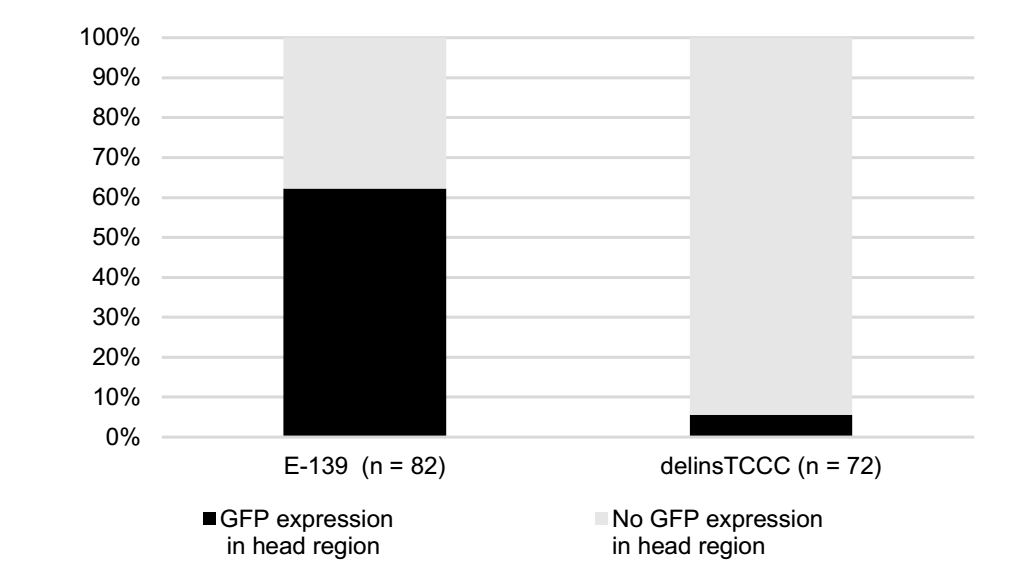

**Supp. Fig. 10. Enhancer element E-139 expression in the brain of zebrafish embryos.** Percentage GFP expression observed in the region of the head of E-139 and delinsTCCC transgenic zebrafish.

**Supplementary Figure 11.**

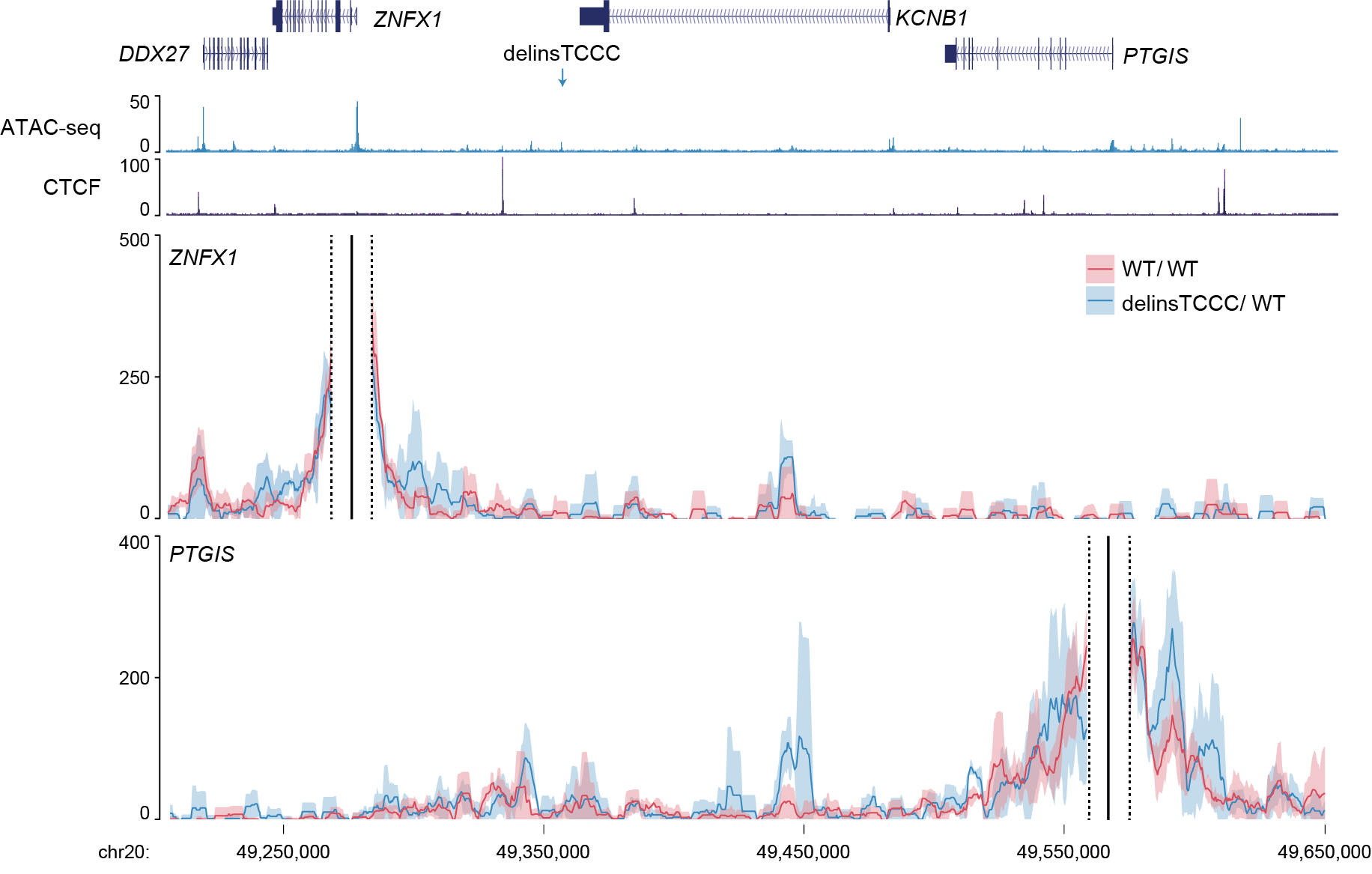

**Supp. Fig. 11. NuTi Capture-C from the *ZNFX1* and *PTGIS* promoters in cardiomyocytes.** Mean interaction count (n=3 independent samples) with one standard deviation for NuTi Capture-C in cardiomyocytes generated from iPSC cells either heterozygous for delinTCCC and uneditied sequence or homozygous unedited. ATAC-seq shows chromatin accessibility in heterozygous cardiomyocytes. CTCF shows ChIP-seq for the CCCTC-binding transcription factor. Region shown chr20:49,210,000-49,650,000 (hg38).

**Supplementary Figure 12.**

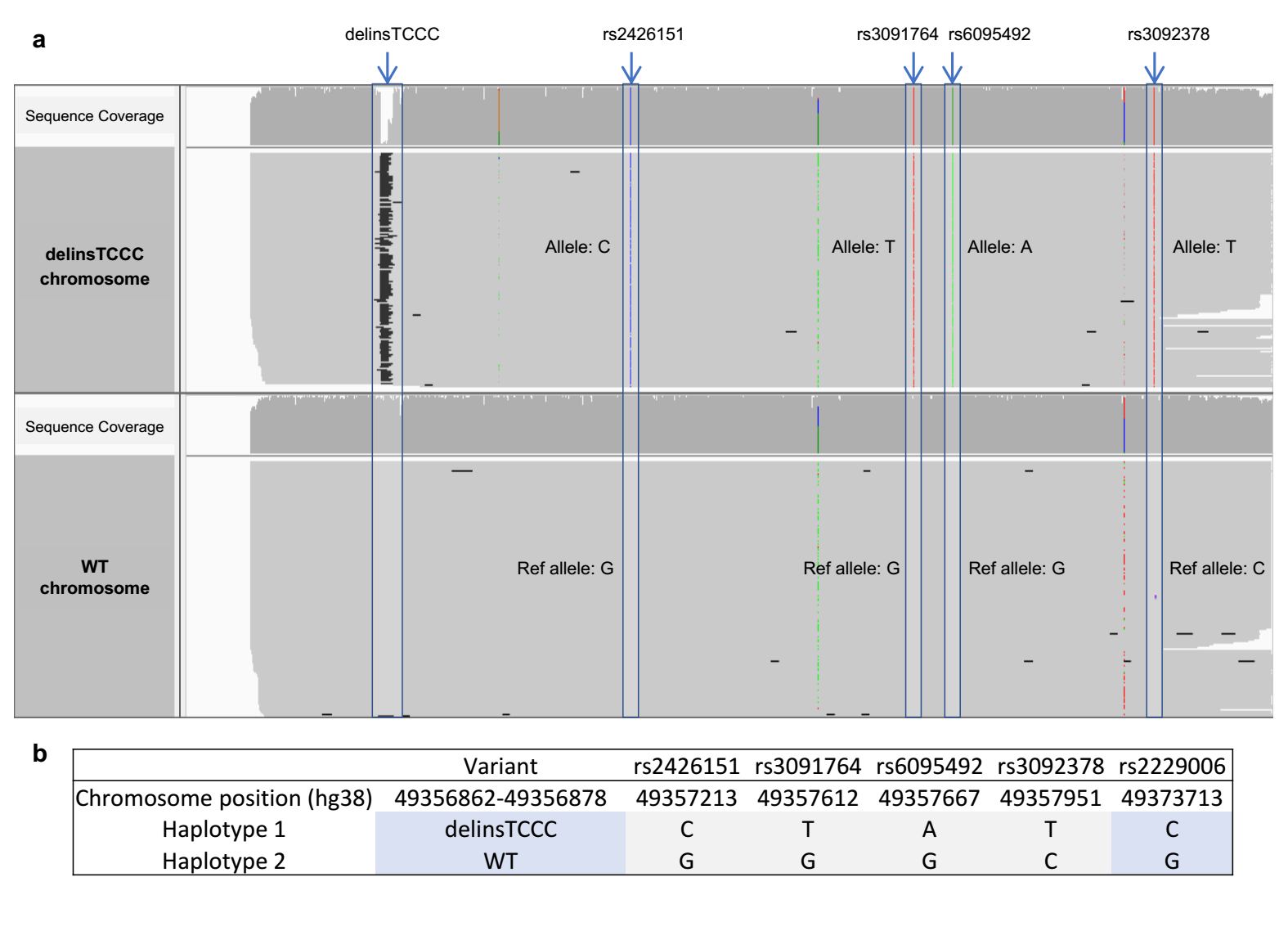

**Supp. Fig. 12. Nanopore sequencing and phasing for the delinsTCCC variant and the rs2229006 SNP. a,** Nanopore sequencing for the heterozygous delinsTCCC C6 iPSC clone identified the genotype of four SNPs (rs2426151, rs3091764, rs6095492, rs3092378) in *cis* with the delinsTCCC variant or WT chromosome. **b**, Using phasing data for the four SNPs and the coding SNP rs2229006, the delinsTCCC variant was determined to be in *cis* with the rs2229006-C allele while the WT chromosome was in *cis* with rs2229006-G allele.

**Supplementary Figure 13.**

**Supp. Fig. 13. Sequence coverage for the individual genomes investigated.** Cumulative distribution of sequence read coverage for 6 genomes investigated from Families 1-3.

**SUPPLEMENTARY TABLES**

**Supplementary Table 1.** Linkage analysis results for Family 1 and Family 2.

| **Family** | **Chr** | **Peak LOD** | **cM start*** | **cM end*** | **bp start** | **bp end** | **size (bp)** |
| --- | --- | --- | --- | --- | --- | --- | --- |
| 1 | 12 | 2.41 | 134.047 | 160.460 | 114,996,664 | 128,399,176 | 13,402,512 |
| 1 | 20 | 2.41 | 62.149 | 76.521 | 43,567,910 | 51,878,414 | 8,310,504 |
| 2 | 20 | 1.52 | 70.294 | 72.872 | 48,212,554 | 50,048,337 | 1,835,783 |

*cM start and end ranges are based on 1 LOD difference from the peak LOD. All coordinates are on build hg38.

**Supplementary Table 2.** Variants shared across six affected individuals from Families 1, 2 and 3.

| Chr | Position (hg38) | Reference allele | Alternate allele | Max_ReMM  score | Gene-level  annotation |
| --- | --- | --- | --- | --- | --- |
| 1 | 25 817 230 | T | TTTGTG | 0.282 | 3_prime_UTR_variant (ENST00000361547.6) |
| 1 | 239 178 250 | C | CAGA | 0.57 | intergenic_region |
| 10 | 131 651 385 | G | GAATA | 0.059 | intergenic_region |
| 11 | 11 376 396 | A | AAACAAAAACAAC | 0.299 | intron_variant (ENST00000227756.4) |
| 11 | 13 094 097 | ACCCTGT | A | 0.45 | intergenic_region |
| 11 | 69 613 144 | A | AAATT | 0.203 | intergenic_region |
| 12 | 64 644 718 | A | ACAAACAAACAAC | 0.087 | intron_variant (ENST00000336061.2) |
| 14 | 58 014 047 | A | AGAAG | 0.511 | intron_variant (ENST00000267485.7) |
| 15 | 93 704 567 | G | GAAAGAAAGAAAC | 0.338 | intergenic_region |
| 16 | 49 154 809 | T | TTAAG | 0.03 | intergenic_region |
| 2 | 17 824 881 | A | AAACC | 0.003 | intergenic_region |
| 2 | 107 123 503 | GC | G | 0.704 | intergenic_region |
| 2 | 168 048 521 | C | CCGGCCCGGCA | 0.336 | intron_variant (ENST00000355999.4) |
| 2 | 240 894 814 | A | ACAAACAAC | 0.214 | 2876bp upstream_gene_variant (ENST00000307486.12) |
| 20 | 49 356 862 | GCCACACCTGTTCCTCT | TCCC | 0.988 | intergenic_region |
| 21 | 20 219 128 | T | TTAGACTG | 0.136 | intergenic_region |
| 22 | 50 589 651 | A | AACAAC | 0.159 | intron_variant (ENST00000463053.1) |
| 4 | 13 472 848 | A | AATAAATAC | 0.149 | intron_variant (ENST00000330852.9) |
| 4 | 42 839 818 | A | ATGAC | 0.152 | intergenic_region |
| 4 | 75 852 159 | T | TAAAG | 0.004 | intergenic_region |
| 4 | 169 135 192 | A | AACAAACAC | 0.49 | intron_variant (ENST00000284637.13) |
| 5 | 24 008 163 | A | ATTTTG | 0.0436289 | intergenic_region |
| 6 | 19 726 125 | T | TAAATAAAC | 0.148 | intergenic_region |
| 7 | 101 131 826 | A | ACAC | 0.767 | intron_variant (ENST00000223095.4) |

**Supplemental Table 3**: Linkage analysis results for Families 1, 2 and combined. Highlighting (in yellow) the interval in which the delinsTCCC was identified.

| **Variant** | **cm** | **pos_b38** | **a1** | **a2** | **SNP#** | **Family 1 LOD** | **Family 1 Risk allele** | **Family 2 LOD** | **Family 2 Risk allele** | **fam3 genotype** | **Combined LOD** |
| --- | --- | --- | --- | --- | --- | --- | --- | --- | --- | --- | --- |
| rs6017575 | 65.47 | 45,461,458 | G | A | 314 | 2.41 | A |  |  |  |  |
| rs2868764 | 65.73 | 46,093,372 | G | T | 315 | 2.41 | T |  |  |  |  |
| rs2425789 | 66.02 | 46,219,177 | G | A | 316 | 2.41 | A |  |  |  |  |
| rs2425874 | 66.44 | 46,335,598 | C | T | 317 | 2.41 | T |  |  |  |  |
| rs429166 | 66.73 | 46,569,632 | G | A | 318 | 2.40 | G |  |  |  |  |
| rs707507 | 67.22 | 46,735,127 | T | C | 319 | 2.40 | T |  |  |  |  |
| rs1333295 | 67.53 | 46,938,867 | G | A | 320 | 2.40 | G |  |  |  |  |
| rs6018248 | 67.97 | 47,057,498 | C | A | 321 | 2.40 | A |  |  |  |  |
| rs6090639 | 68.18 | 47,206,364 | C | T | 322 | 2.41 | T |  |  |  |  |
| rs6066318 | 68.56 | 47,422,521 | G | T | 323 | 2.41 | T |  |  | T/G |  |
| rs2235734 | 68.82 | 47,663,405 | T | G | 324 | 2.41 | G |  |  | G/G |  |
| rs6012271 | 69.24 | 47,805,339 | G | A | 325 | 2.41 | A |  |  | A/A |  |
| rs2426053 | 69.54 | 47,938,041 | A | C | 326 | 2.41 | C |  |  | C/C |  |
| rs13044311 | 69.99 | 48,187,136 | T | C | 327 | 2.41 | T |  |  | T/C |  |
| rs852358 | 70.18 | 48,208,229 | T | C | 328 | 2.41 | C |  |  | C/T |  |
| rs391144 | 70.49 | 48,238,264 | G | A | 329 | 2.41 | A |  |  | G/A | 3.34 |
| rs432379 | 70.61 | 48,318,896 | C | T | 330 | 2.41 | T |  |  | C/T | 3.50 |
| rs1555581 | 70.84 | 48,404,632 | A | C | 331 | 2.41 | A |  |  | C/A | 3.86 |
| rs6019278 | 71.02 | 48,548,239 | T | C | 332 | 2.41 | C |  |  | T/C | 4.02 |
| rs6019300 | 71.28 | 48,584,971 | G | A | 333 | 2.41 | A | 1.40 | A |  | 4.18 |
| rs11699576 | 71.46 | 48,733,341 | C | A | 334 | 2.41 | A | 1.48 | A |  | 4.26 |
| rs4141783 | 71.69 | 49,162,065 | A | G | 335 | 2.41 | G | 1.49 | G | G/G | 4.27 |
| rs6019687 | 71.70 | 49,215,367 | G | T | 336 | 2.41 | T | 1.49 | T |  | 4.27 |
| rs2273145 | 71.70 | 49,223,054 | T | C | 337 | 2.41 | C | 1.49 | C |  | 4.27 |
| rs674063 | 71.73 | 49,333,298 | T | C | 338 | 2.41 | T | 1.49 | T | C/T | 4.27 |
| rs237450 | 71.73 | 49,365,591 | G | A | 339 | 2.41 | G | 1.49 | G | A/G | 4.27 |
| rs237480 | 71.74 | 49,377,361 | A | C | 340 | 2.41 | C | 1.48 | C |  | 4.26 |
| rs237475 | 71.80 | 49,432,969 | T | C | 341 | 2.41 | T | 1.43 | T | C/T | 4.21 |
| rs500670 | 72.30 | 49,591,123 | A | C | 342 | 2.41 | C |  |  | C/A | 3.83 |
| rs4297943 | 72.59 | 49,807,219 | A | G | 343 | 2.41 | G |  |  | A/G | 3.59 |
| rs6125878 | 72.88 | 50,048,337 | G | T | 344 | 2.41 | T |  |  |  | 3.45 |
| rs6020394 | 73.04 | 50,250,779 | C | T | 345 | 2.41 | T |  |  | T/C | 3.35 |
| rs768175 | 73.52 | 50,621,890 | A | C | 346 | 2.41 | C |  |  | C/C | 3.24 |
| rs6067559 | 73.91 | 50,815,866 | T | C | 347 | 2.41 | C |  |  | C/C |  |
| rs6020905 | 74.32 | 51,007,608 | C | A | 348 | 2.41 | A |  |  | A/A |  |
| rs170717 | 74.74 | 51,122,227 | A | G | 349 | 2.41 | G |  |  | G/A |  |
| rs6021046 | 74.95 | 51,209,643 | T | C | 350 | 2.41 | C |  |  | C/C |  |
| rs6067728 | 75.12 | 51,296,680 | G | A | 351 | 2.41 | A |  |  | A/A |  |
| rs2426311 | 75.61 | 51,429,660 | C | T | 352 | 2.41 | T |  |  | T/T |  |
| rs6021247 | 75.88 | 51,492,442 | A | G | 353 | 2.41 | G |  |  | G/G |  |
| rs926668 | 76.00 | 51,544,919 | T | C | 354 | 2.41 | C |  |  | C/T |  |
| rs2151119 | 76.20 | 51,750,432 | C | T | 355 | 2.41 | T |  |  | T/C |  |

**Supplementary Table 4.** GFP expression in the heart of transgenic zebrafish**.**

| **Transgenic fish** | **Strong (n)** | **Weak (n)** | **None (n)** |
| --- | --- | --- | --- |
| E-139 (n =286) | 152 | 67 | 67 |
| WT (n = 169) | 19 | 84 | 66 |
| delinsTCCC (n = 160) | 104 | 34 | 22 |
| Neg Control (n = 156) | 0 | 13 | 143 |

n= number of transgenic zebrafish embryos per category

**Supplementary Table 5.** Allele specific expression using RNA sequencing.

|  | Total reads | G allele | C allele | G allele % | C allele % | Fold change |
| --- | --- | --- | --- | --- | --- | --- |
| Differentiation A | 11 | 9 | 2 | 82 | 18 | 4.5 |
| Differentiation B | 15 | 13 | 2 | 87 | 13 | 6.5 |
| Differentiation C* | 6 | 2 | 4 | 33 | 67 | 0.5 |
| Mean | 10.7 | 8.0 | 2.67 | 67 | 33 | 3.8 |

* Low number of reads for this sample could explain differences in allelic expression.

**Supplementary Table 6.** Summary quality metrics for WGS

| **Individual** | **M reads mapped** | **Percentage of bases covered >= 10X**  **(whole genome)** | **Percentage of bases covered >= 10X**  **(exonic regions)** | **Median coverage** | **Duplicated reads (%)** | **Mean read length** |
| --- | --- | --- | --- | --- | --- | --- |
| F1: II5 | 849 | 92.32 | 98.00 | 26.0X | 7.15 | 100 bp |
| F1: III3 | 860.7 | 92.36 | 98.00 | 26.0X | 1.77 | 100 bp |
| F1: IV1 | 874.3 | 93.15 | 99.30 | 35.0X | 9.34 | 148 bp |
| F1: II1 | 877 | 93.75 | 99.70 | 37.0X | 9.97 | 148 bp |
| F2: IV1 | 978.3 | 93.81 | 99.80 | 39.0X | 14.23 | 148 bp |
| F3: II1 | 823.3 | 92.26 | 99.10 | 35.0X | 16.48 | 150 bp |

F1 = Family 1; F3 = Family 3

**Supplementary Table 7.** Chromosome conformation capture oligonucleotides

| **Name** | **Chromosome 20 coordinates (hg38)** | **Pool** | **Sequence** |
| --- | --- | --- | --- |
| ZFX1_L | 49277559-49277629 | Promoter | GATCGAAGTGCTGAGGGCAGAAGCGGTGACGGTGTCGGGGGTGCTGGGGAGAGCGACGGGCCAGCTGAGG |
| ZFX1_R | 49278910-49278980 | Promoter | GCCTTCTGCACGGGCCTGAGAAGCCCTTGGCTGGTGTAAATGATGACTTCACTTTTTTCCCCATCAGATC |
| PTGIS_L | 49568413-49568483 | Promoter | GATCAAGGGGAAAGGAGATAGATGGGGGACTGAGGGGGATGTGGTGGACTGGGGGGTGATGGAAGGGAGG |
| PTGIS_R | 49569446-49569516 | Promoter | CTCCTGGTCTCCTGGCTTCCATCCTTATCAGCCAGGGTCCTGACAAGGCCCTTCGAGAACCAGCATGATC |
| KCNB1_L_L | 49484231-49484301 | Promoter | GATCAACAAGTCGGGGCTGAGCTTCGGCTCCGCGCTCGCTGCTATTCTGTCCCGCACTTCGGAGCGCCCG |
| KCNB1_L_R | 49484342-49484412 | Promoter | AGCGAGCGCGGCTTAACTCCTGCCTGCCCGGCCCAGCCTGCCGGGGAGGCCGGGGGCGGGCTCCGGGATC |
| KCNB1_S_L | 49482485-49482455 | Promoter | GATCCGGCCGCCCCCGCCCCCCCTGCCCCCCCAGGCCGCTGTCACTCGACGGCAAGGCCCGCTGCTGCGG |
| KCNB1_S_R | 49482933-49483003 | Promoter | GACCCAGCCGCCCGGGGCGCTGTCCGCCCCTTAGAGTGCTGAAACTCTCTTCATTATGGGTCTCCCGATC |
| INDEL_L | 49356807-49356877 | Enhancer | GATCATCAGGAGGGATAGGTCCTCAAACAGGCGATGAGCTCAGGACATGGGCATTGCCACACCTGTTCCT |
| INDEL_R | 49356933-49357003 | Enhancer | TGAATGTGATGTGCATACTTGTGCATGGGCAGGAGAGCAGACAGGGACCAAGCCAAGGCTGACAGGGATC |
| E-163_R | 49321069-49321139 | Enhancer | CGGAGTGGGGGCTGGAGGAACCAGGAGCCTCAAAGCTTTGCTTTCACCTTTGTTTTGCTCTTTTGGGATC |
| E-139_L | 49345180-49345250 | Enhancer | GATCTCATCCCTGAACACTGAGGAAGAAAGGGTGGGGGTGAAGCTGCTCGCTGGCAGCATCTGACATAGT |
| E-139_R | 49345666-49345736 | Enhancer | TTTGTCACTATTGCATTACATAAGAGGAAAGACATTTTTAAAGAATGCGGCTACATTTAGTAATTGGATC |
| E-98_L | 49385395-49385465 | Enhancer | GATCAATTCAAGTTCACCAACATCTGCGCATCTGTTCCATTCTCTCCACCTGCACAGTCCCCACCCTCCC |
| E-98_R | 49385692-49385762 | Enhancer | TGTCTCCTTTTCCTCCTTGCTGGCTGGCAGGCAGATGTGCTGGGCAGAGCTGGAGCAACTGTCTTGGATC |
| E-39_L | 49444906-49444976 | Enhancer | GATCTCTAACCCTAGTCATACACTGAGTTCAGCCCTGGCCTCAGATGCCCAGCCATCTCCAATTGCTGCT |
| E-39_R | 49445957-49446027 | Enhancer | GTTAGGAGAGTCAAGAGTCAAGTATCCAGTGAGCTGGTCATTCGGATACCGAGGTGGGAAACAGGTGATC |

**Supplementary Table 8.** Enhancer sequences for transgenic zebrafish

| Transgenic zebrafish | Sequence |
| --- | --- |
| WT human KCNB1  +125 enhancer | CAAACTCCTTTGCACTCGGGAGACTGGAGAGTAGATTCCTTTAGAAGTTT GTTCTTGTGATTCACGTTCCCTCTCCCAGGCTATCCCCTTCAATAGCTAG GATTGGGAATAAGAATGCACACATGTACAGAAAAGCACAGAAACATCTGC CCTGCCTTAGTTCCCTACAGGGTCTCATCAGAGGTGGTTTCATTAGCCCC AACCCTTTGCAGCCTGGCACCTTCAGACATGCCACCAGCTGGCTCTCCTA ATATTTGCTGTGGGGCCATGTGATCATCAGGAGGGATAGGTCCTCAAACA GGCGATGAGCTCAGGACATGGGCATTGCCACACCTGTTCCTCTCCCCAAG GAGGGGGGAAAGCTGTGCAGTCACAGACAACGTGGCCTTTTATAAGCTTG AATGTGATGTGCATACTTGTGCATGGGCAGGAGAGCAGACAGGGACCAAG CCAAGGCTGACAGGGATCAGAGGGAAGGACAGGGGTGTTTGCGGGTGCCT GCAATGTGGCAGTGGCTGAGCTGGGCCTGTGTGCCTGTACACCATTGTGt ttcagatatttttgactgtcaagttgcaggaagaagtatattttgcagca tgacctgtaacacacacacatgcacacacaattgacatgaaagtttcatg aatctgtacttaccttttatgagcaatgcatgctgactttgattcgattc |
| Mutant human KCNB1  +125 enhancer | CAAACTCCTTTGCACTCGGGAGACTGGAGAGTAGATTCCTTTAGAAGTTT GTTCTTGTGATTCACGTTCCCTCTCCCAGGCTATCCCCTTCAATAGCTAG GATTGGGAATAAGAATGCACACATGTACAGAAAAGCACAGAAACATCTGC CCTGCCTTAGTTCCCTACAGGGTCTCATCAGAGGTGGTTTCATTAGCCCC AACCCTTTGCAGCCTGGCACCTTCAGACATGCCACCAGCTGGCTCTCCTA ATATTTGCTGTGGGGCCATGTGATCATCAGGAGGGATAGGTCCTCAAACA GGCGATGAGCTCAGGACATGGGCATTtcccCCCCAAG GAGGGGGGAAAGCTGTGCAGTCACAGACAACGTGGCCTTTTATAAGCTTG AATGTGATGTGCATACTTGTGCATGGGCAGGAGAGCAGACAGGGACCAAG CCAAGGCTGACAGGGATCAGAGGGAAGGACAGGGGTGTTTGCGGGTGCCT GCAATGTGGCAGTGGCTGAGCTGGGCCTGTGTGCCTGTACACCATTGTGt ttcagatatttttgactgtcaagttgcaggaagaagtatattttgcagca tgacctgtaacacacacacatgcacacacaattgacatgaaagtttcatg aatctgtacttaccttttatgagcaatgcatgctgactttgattcgattc |
| Negative control | AAAAATGTAGAAGTTAATGTATTAtttctgcatatcggtctgtggactaa gctttatacgatgcattcattctctttatttacctcaaatgtgggaagta gctattattgcctcttcctgacaacaacaaaaaaagcctgaggactggag atgggctgtgccttgcccaaggtcacatggtgaattagtgccagaactga gactccactgggtgcagtggcccacgcctataatctcagtactttgagag gctgaggcaggtagatcgcttgagcccaggagtttgagaccagcctgggc aacatggtgagaccctgtttctactaaaataaaaaatacaacagttagct aggcatggtggtgtgcaatgtagtcccagctacttgggaagctgaggcag gagaatcacatgaacctgggtgatggaggttgcagtgagccgagatcgag ccactgcactccagcctgggtgacacagcgagactctatcacagacaaac aaacagacagacaaagaattgagactctactccagGCCTTCTAACCTGTC CAGGGGGTCTCCACTTCTCCAGGTACCTCTTCCTTTACTGGCGGAAGCCA TGCCGAGGTGTGGACAGAGGGGTGGTGATACCTCTGTAGTTATCACCtac Agtagggtggtcacacgtcacactggtgttacacaatctgggctcaaatc |
| E139 | TCCCGGCCAGGGTTTGACGTGAAAAGTTTGGGGAATGGTAATAGCACGGG TCACGTTGACTTGTGAGCAATGGGCAAAAGATACCTGGTTTGGGCAGAGG GGAAAAATGCCTTGGGCCTCCCAAAAGCCCAGGAGGCTGGAGCCTCCTCA ATCTATTTACCAAGACTTTTTTATTTTTTTTTTCAAAATTGTACATTGTC TTCGAGAGAGCGGACCGTCTCTCCCTGCCTCGGGAGGGAAGCGGGTTAGT CATAAACTATGTCAGATGCTGCCAGCGAGCAGCTTCACCCCCACCCTTTC TTCCTCAGTGTTCAGGGATGAGATCATTTCATTTGGAGCCAATTATAGAC GGAGAATGCTGGGGATGTGCAAAGAAACCTCCTCAGAACCCTTCCCTCCC ACATCACAGGCTGTACCCTGCCAGGATGGGCAAGAACAAGAGGCCAGGCG CCCAGCCCCAGGTTCGACCCCAGGGTTTCTGAAGGCCATGCATTCCTAAG CCTTAGCTTGGACCTGTCAAGCTAGGATGCCAGGCTGTTGGCTAGACAAA CCCTGTTGGCCTCAAAAGTTCAGGGCTGTATGGGCCCTTAAAGATCATCG CTGTTCAGATGAGGAAACTGAGGCTCAGAGAAGGG |
